# Epidemiological characteristics of three SARS-CoV-2 variants of concern and implications for future COVID-19 pandemic outcomes

**DOI:** 10.1101/2021.05.19.21257476

**Authors:** Wan Yang, Jeffrey Shaman

## Abstract

Three SARS-CoV-2 variants classified as variants of concern – B.1.1.7, B.1.351, and P.1 – have spread globally. To characterize their viral and epidemiological properties in support of public health planning, we develop and apply a model-inference system to estimate the changes in transmissibility and immune escape for each variant, based on case and mortality data from the country where each variant emerged. Accounting for under-detection of infection, disease seasonality, concurrent non-pharmaceutical interventions, and mass-vaccination, we estimate that B.1.1.7 has a 46.6% (95% CI: 32.3 – 54.6%) increase in transmissibility but nominal immune escape from protection induced by prior wild-type infection; B.1.351 has a 32.4% (95% CI: 14.6 – 48.0%) increase in transmissibility and 61.3% (95% CI: 42.6 – 85.8%) immune escape; and P.1 has a 43.3% (95% CI: 30.3 – 65.3%) increase in transmissibility and 52.5% (95% CI: 0 – 75.8%) immune escape. Model simulations indicate that B.1.351 and P.1 could supplant B.1.1.7 dominance and lead to increased infections. Our findings highlight the importance of preventing the spread of B.1.351 and P.1, in addition to B.1.1.7, via continued preventive measures, prompt mass-vaccination of all populations, continued monitoring of vaccine efficacy, and possible updating of vaccine formulations to ensure high efficacy.

## Main text

Multiple SARS-CoV-2 variants have been identified since summer 2020. Among these, three variants – namely, B.1.1.7, B.1.351, and P.1 – have been classified as variants of concern (VOCs), per evidence indicating these genotypes are substantially more transmissible, evade prior immunity (either vaccine-induced or conferred by natural infection with wild-type virus), increase disease severity, reduce the effectiveness of treatments or vaccines, or cause diagnostic detection failures.^1, 2^ Multiple lines of evidence indicate the B.1.1.7 variant is roughly 50% more transmissible than wild-type virus but does not produce antigenic escape.^3–6^ Further, several studies have shown that both the B.1.351 and P.1 variants are resistant to neutralization by convalescent plasma from individuals previously infected by wild-type SARS-CoV-2 viruses and sera from vaccinated individuals;^5–9^ however, changes to the transmissibility of these latter two variants are less well resolved. A better understanding of the transmissibility and immune escape properties of VOCs – in particular, B.1.351 and P.1 at present – is needed to anticipate future COVID-19 pandemic outcomes and support public health planning.

In this study, we develop a model-inference system to estimate the relative change in transmissibility and level of immune evasion for different SARS-CoV-2 variants, while accounting for under-detection of infection, delays of reporting, disease seasonality, non-pharmaceutical interventions (NPIs), and vaccination. Testing using model-generated synthetic incidence and mortality data indicates this inference system is able to accurately identify shifts in transmissibility and immune evasion. We then apply the validated inference system in conjunction with incidence and mortality data from the UK, South Africa, and Brazil – the three countries where VOCs B.1.1.7, B.1.351, and P.1 were first identified, respectively – to estimate the change of transmissibility and immune evasion for the three VOCs, separately. We further use these inferred findings in a multi-variant, age-structured model to simulate epidemic outcomes in a municipality like New York City (NYC) where multiple variants, including B.1.1.7, B.1.351, and P.1, have been detected.

### The model-inference system and validation

We first tested our model-inference system using 10 model-generated synthetic datasets, depicting different combinations of population susceptibility prior to the emergence of a new variant, changes in transmissibility and immune evasion for the new variant, and infection-detection rate. As population susceptibility, interventions, and disease seasonality can all affect apparent transmissibility at a given time and in order to focus on variant-specific properties, here we defined transmissibility as the average number of secondary infections per primary infection, *after removing the effects of these three factors* (see Methods). We then quantified the change in transmissibility as its relative increase once the new variant becomes dominant. Similarly, we quantify the level of immune evasion as the increase in susceptibility after the new variant becomes dominant, relative to prior population immunity from wild-type infection.

Fig. 1 shows example test results comparing model-inference system estimates with model-generated ‘true’ values of transmissibility and susceptibility using an infection-detection rate of 20%. Across a range of epidemic dynamics, the model-inference system is able to fit both weekly incidence and mortality data (Fig. 1A) and estimate the transmissibility and susceptibility over time for both the initial pandemic wave and the subsequent pandemic wave caused by a new variant (Fig. 1B). In addition, when aggregated over both pandemic waves, the model-inference system is able to estimate the relative changes in transmissibility and immune evasion due to a new variant (Fig. 1C). When the system is less well constrained (e.g., an infection-detection rate of 10%), model estimates, albeit less accurate, still closely track true values in most instances (Fig S1).

**Fig 1.**
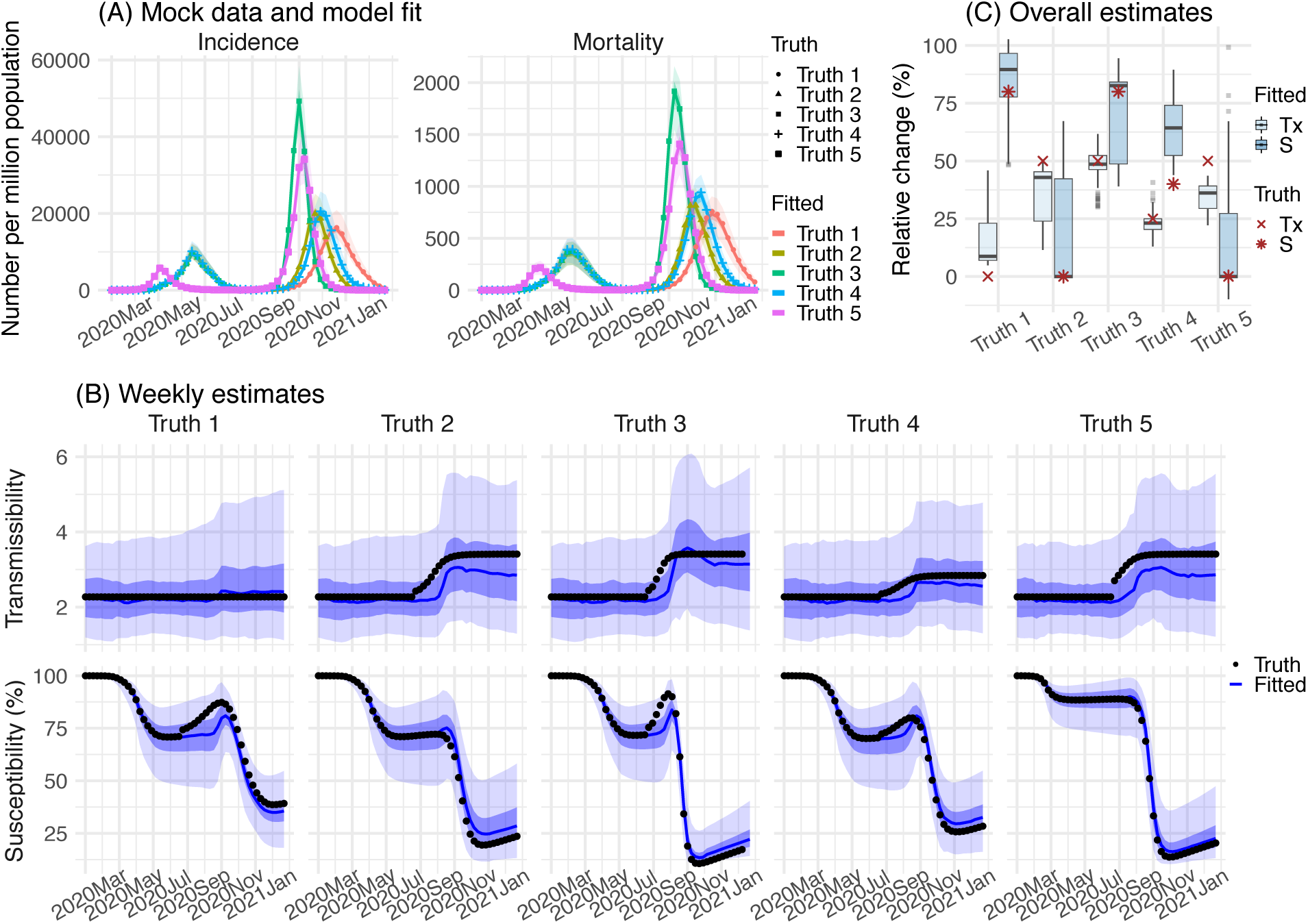
Model-inference system validation using model-generated synthetic data with an infection-detection rate of 20%. (A) 5 sets of synthetic data (dots) and model-fits to each dataset; lines show mean estimates and surrounding areas show 50% (dark) and 95% (light) CrIs. (B) weekly model estimated transmissibility and population susceptibility; lines show mean estimates and surrounding areas show 50% (dark) and 95% (light) CrIs, compared to the true values (dots). (C) overall estimates of the change in transmissibility and immune evasion (boxes and whiskers show model estimated median, interquartile range, and 95% CI from 100 model-inference simulations) compared to the true values (dots).

### Reconstructed pandemic dynamics in the three countries

Following the initial emergence of SARS-CoV-2 in early 2020, the UK, South Africa, and Brazil experienced very different epidemics (Fig. 2). The model-inference system is able to recreate the observed incidence and mortality epidemic curves for all three countries (Fig 2 A, C, and E). Further, cross-validation using independent data shows that the model estimates closely match measured SARS-CoV-2 prevalence in the UK^10, 11^ and serology measures of cumulative infection rates in South Africa^12, 13^ and Brazil,^14^ respectively (Fig 2 B, D, and F). These results indicate the model-inference system accurately estimates the underlying transmission dynamics for all three countries.

**Fig 2.**
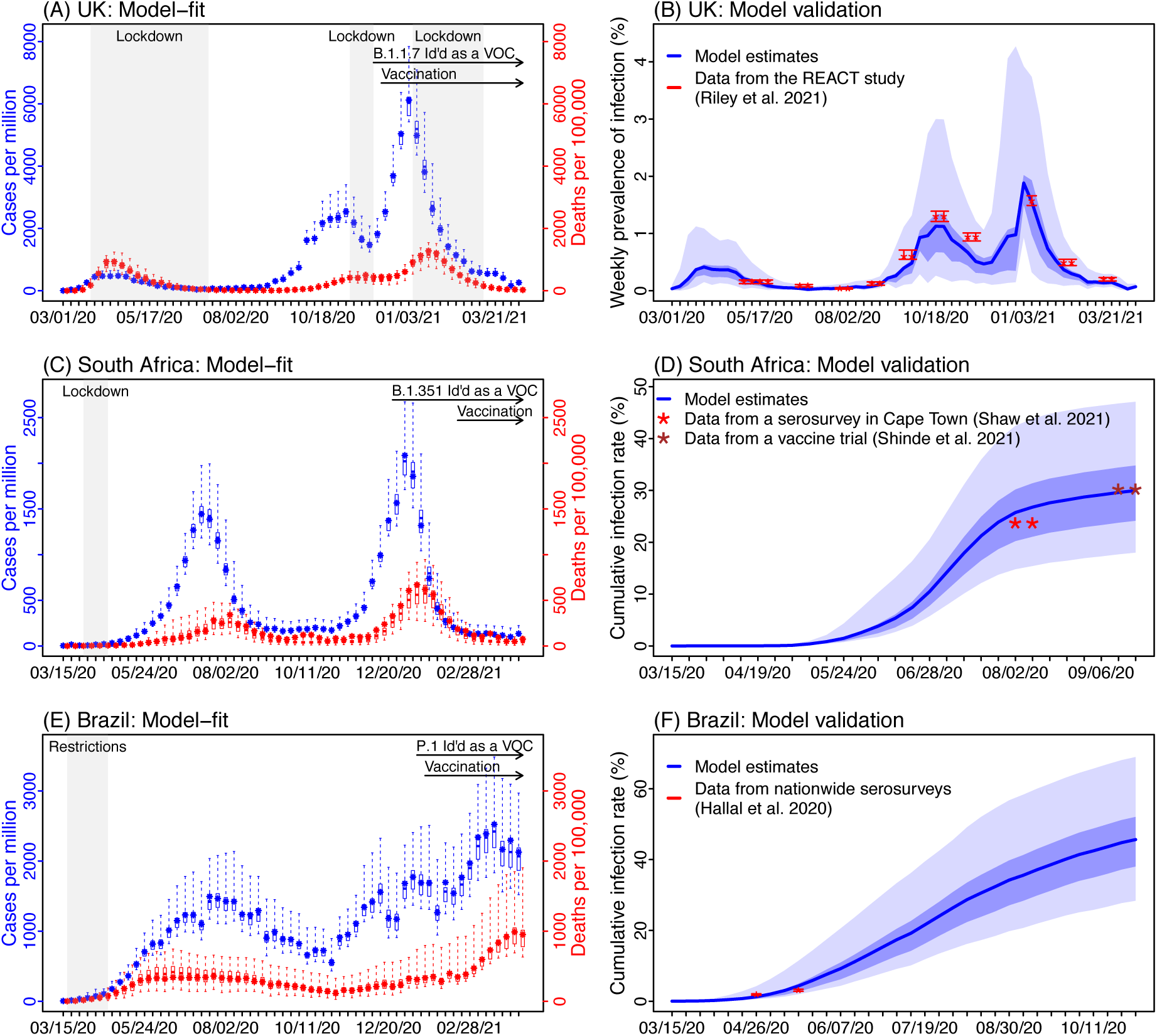
Model-inference system fit to data for the three countries and validation using independent datasets. The left column shows the model fit to reported weekly case and mortality data for the UK (A), South Africa (C), and Brazil (D). Dots show the weekly number of cases (in blue) and deaths (in red) per 1 million persons; boxes and whiskers show corresponding model estimates (mean, 50% and 95% CrIs). Grey shaded boxes indicate the timing of lockdowns or key periods of restricted activity; horizontal arrows indicate the timing of variant identification and vaccination rollout. The right column compares available, independent measurements to corresponding model estimates. Red dots and error bars show measured prevalence over 10 periods of time from the REACT study for the UK (B), cumulative infection rates from two serology studies in South Africa (D), and cumulative infection rates from two nationwide household serosurveys in Brazil (F). Blue lines and surrounding areas show model estimated mean, 50% (dark) and 95% (light) CrIs.

In the UK, a prompt lockdown allowed the country to contain the first pandemic wave (Fig. 3). The real-time effective reproduction number (*R_t_*), which measures the average number of secondary infections at a given point in time, dropped from 2.2 (95% CrI: 1.0 – 3.9) during the week of 3/1/2020 to below 1 during the week of 3/22/2020, the first week of the lockdown (Fig 3A). By the week of 6/28/2020 (the week with the lowest incidence following the first pandemic wave), 6.4% (95% CrI: 3.6 – 12.3%) of the UK population are estimated to have been infected. However, with the relaxation of intervention measures during the summer, transmission gradually increased again (as indicated by the estimated *R_t_* >1), leading to a large surge of infections in the autumn of 2020 (Figs 2A-B and 3A). A second lockdown implemented in Nov 2020 reduced transmission transiently (*R_t_* was below 1 during the 4-week lockdown period; Fig 3A). Shortly thereafter widespread transmission of the B.1.1.7 variant led to a further increase of cases before this activity was curtailed by a third lockdown and mass-vaccination.

**Fig 3.**
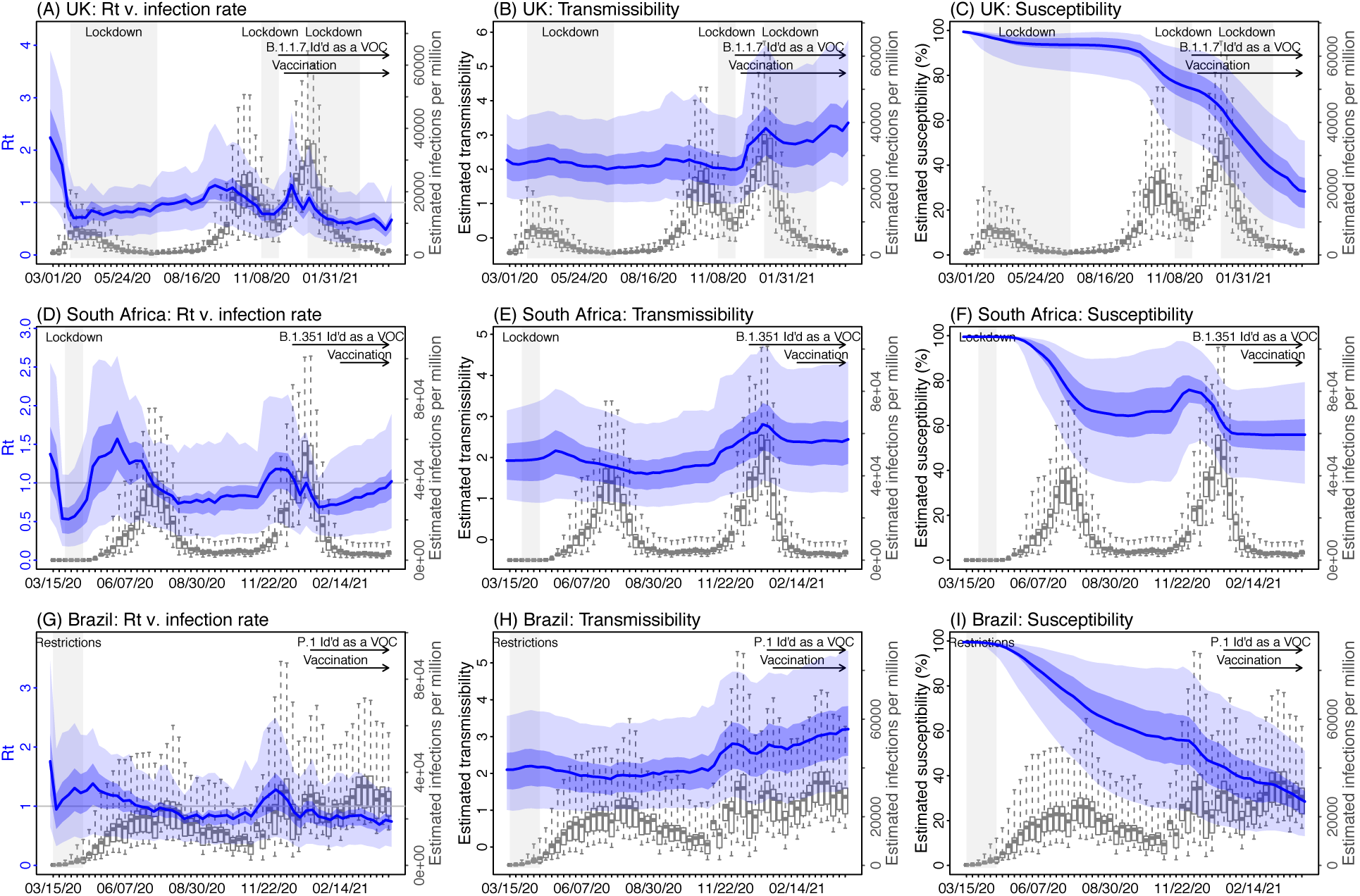
Key model-inference system estimates. Left column shows estimated real-time reproduction number *R_t_*, middle column shows estimated transmissibility, and right column shows estimated population susceptibility for each week during the study period, for the three countries. For comparison, estimated weekly infection rates are superimposed in each plot (right y-axis). Blue lines and surrounding areas show the estimated mean, 50% (dark) and 95% (light) CrIs. Boxes and whiskers show the estimated weekly infection rates (mean, 50% and 95% CrIs). Grey shaded areas indicate the timing of lockdowns or key periods of restricted activity; horizontal arrows indicate the timing of variant identification and vaccination rollout. *Note that the transmissibility estimates (middle column) have removed the effects of changing population susceptibility, NPIs, and disease seasonality; thus, the trends are more stable than the reproduction number (R_t_; left column) and reflect changes in variant-specific properties*.

In South Africa, initial transmission was low likely due to a strong public health response (a lockdown was implemented from 3/26/20 to 4/16/20) and less conducive conditions for transmission during southern hemisphere summer and autumn (Figs 2C, 3D, and S2B). However, as the country relaxed intervention measures and entered the winter, transmission increased substantially from May 2020 onwards, leading to a large pandemic wave during May – Sep 2020. After accounting for under-detection of infection (Fig S3C), consistent with serology data,^12, 13^ the model-inference system estimates that 30.0% (95% CrI: 18.0 – 47.1%) of the population had been infected by the week of 9/20/2020 (i.e., the week with the lowest incidence following the first pandemic wave; Fig 2D). After two months with relatively low incidence, the emergence of the B.1.351 variant led to a resurgence of infections in late 2020 and a larger second pandemic wave (Fig 3D). By the week of 4/11/21, another 38.6% (95% CrI: 24.1 – 61.1%) of the population are estimated to have been infected, including re-infections.

In Brazil, no national lockdown was implemented during the pandemic. A large first pandemic wave occurred during Mar – Oct 2020 (Figs 2E and 3G). By the week of 11/1/2020 (i.e., the week with the lowest incidence following the first wave), 45.7% (95% CrI: 28.4 – 69.0%) of the population are estimated to have been infected (Fig 3F). This estimate includes all infections and thus is much higher than the reported number of cases (3.77% of the population; see estimated infection-detection rates in Fig S3E). In addition, unlike the UK and South Africa where the pandemic wave rose and fell quickly, pandemic activity – based on national incidence and mortality curves – remained at high levels for a much longer duration (Fig. 2E). This may be due to the larger geographical area of Brazil, the aggregative nature of country-level incidence and mortality data combining multiple outbreak waves from different sub-regions of the country, and the lack of national restrictions to curb the pandemic. Despite this large first pandemic wave, the emergence of the P.1 variant led to a second large pandemic wave from Dec 2020 onwards. Similar traveling waves through the country were evident from the incidence curve (Fig 2E). By the week of 4/11/21, an additional 60.7% (95% CrI: 40.5 – 92.0%) of the population are estimated to have been infected, including re-infections.

### Estimated increased transmissibility and immune evasion

Accounting for concurrent NPIs, vaccination and seasonal transmission trends (Fig S2), model-inference system estimates also enable assessment of key properties specific to the three variants. Estimated transmissibility increased in conjunction with the widespread presence of the new variant in each country (Fig 3B for B.1.1.7, 3E for B.1.351, and 3H for P.1). Overall, estimated viral transmissibility increased by 46.6% (95% CI: 32.3 – 54.5%) for the B.1.1.7 variant, 32.4% (95% CI: 14.6 – 48.0%) for the B.1.351 variant, and 43.3% (95% CI: 30.3 – 65.3%) for the P.1 variant, compared to the wild-type virus (Table 1). In addition, the model-inference system also detects large increases of population susceptibility for the B.1.351 and P.1 variants, but not the B.1.1.7 variant (Fig. 3F and 3I vs. 3C; and Table 1). Specifically, the model estimates immune evasion for the B.1.351 variant among 61.3% (95% CI: 42.6 – 85.8%) of the population infected with the wild-type virus during the first wave in South Africa and for the P.1 variant among 52.5% (95% CI: 0 – 75.8%) of the population infected with the wild-type virus during the first wave in Brazil.

**Table 1.** Estimated changes in transmissibility and level of immune evasion. Numbers show model estimated mean (95% CI) from 100 model-inference runs.

| Location | Variant | Changes in |  |
| --- | --- | --- | --- |
|  |  | Transmissibility (%) | Immune evasion (%) |
| United Kingdom | B.1.1.7 | 46.6 (32.3, 54.6) | 3.9 (0, 36.2) |
| South Africa | B.1.351 | 32.4 (14.6, 48) | 61.3 (42.6, 85.8) |
| Brazil | P.1 | 43.3 (30.3, 65.3) | 52.5 (0, 75.8) |

### Competition among variants and potential future outcomes

As many places have now detected one or more of the three VOCs locally, it is important to understand the potential pandemic outcomes given the characteristics of and competition among variants, interactions with ongoing NPIs, and mass-vaccination. We thus use a multi-variant, age-structured model to simulate potential pandemic outcomes for the period from May 2021 to Aug 2021 under scenarios with different variant prevalence, NPIs, and vaccine efficacy. We focus on NYC for which detailed data and estimates (e.g., contact patterns, variant prevalence, and vaccination rates) are available.

Fig 4A and Fig S4 show example projections of infections and mortality assuming a best-case scenario in which vaccine-induced immunity is as effective against all three VOCs as for the wild-type virus. At the time of these simulations (i.e., end of April 2021), the B.1.1.7 variant was the predominant VOC in NYC; however, given their estimated propensity for immune escape, both B.1.351 and P.1 could outcompete B.1.1.7 and become predominant in the coming months (Fig 4A, top panel). The relative prevalence of B.1.351 and P.1 depends largely on their initial introduction and establishment in the population. These two variants would arise at similar rates and co-dominate, if they are introduced and established in the population simultaneously (Fig 4A, left panel). However, should either be established in the population ahead of the other, it would become dominant and suppress but not preclude the rise of the other variant (Fig 4A, middle and right panels). In addition, the B.1.351 variant would be slightly more competitive if the vaccines are less effective against it than the P.1. variant (Fig 4B and Table S1), as has been shown in laboratory studies^7, 9^.

**Fig 4.**
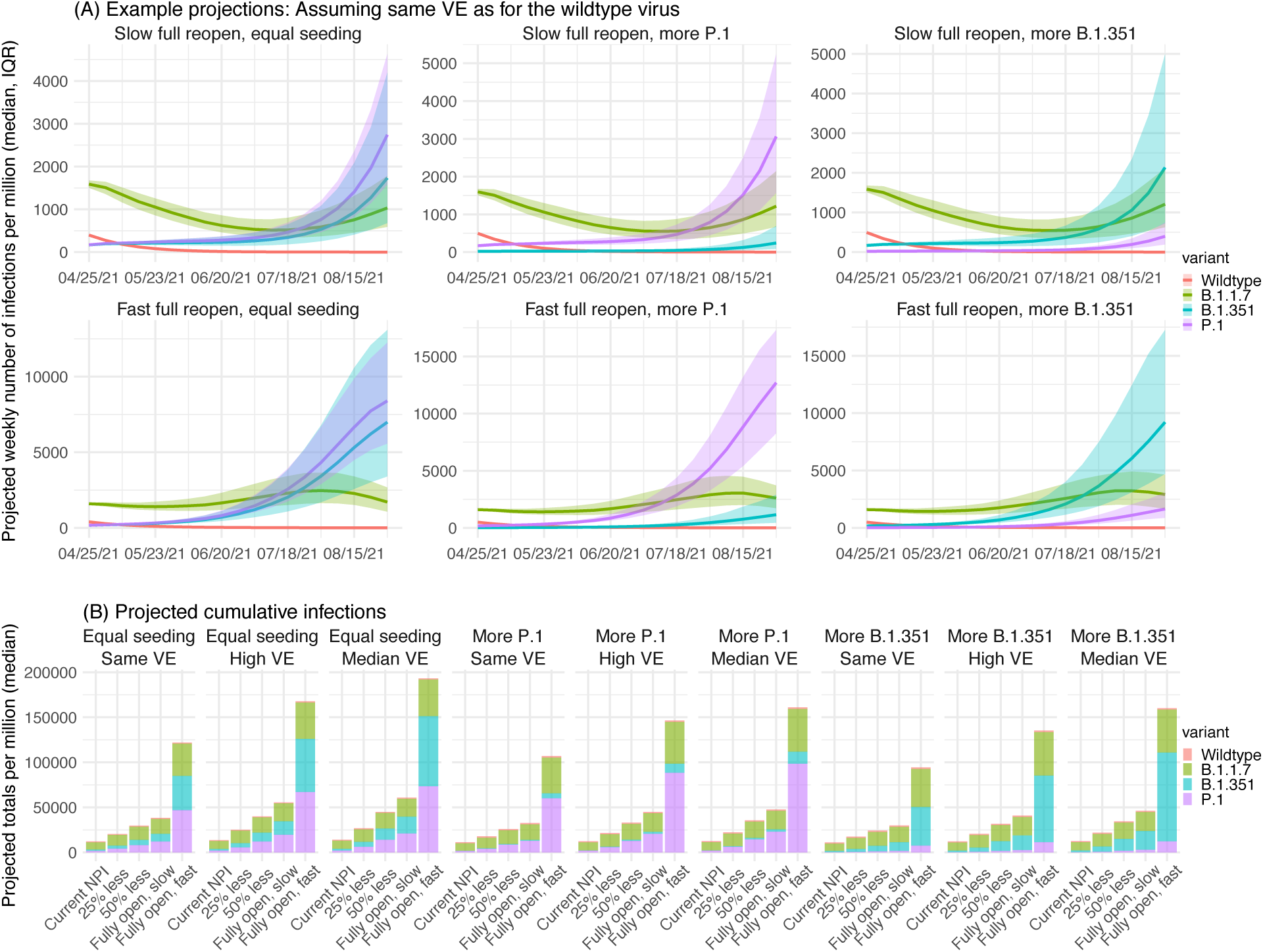
Model projections of infection, under different scenarios of VOC co-circulation, NPIs, and vaccine efficacy (VE). (A) Example projected epidemic trajectories for each variant assuming the VE is as high as for the wild-type virus. Lines and surrounding areas show model projected median and interquartile range (color-coded for each variant as indicated by the legend). (B) Tallies over the entire simulation period (May – Aug 2021) for different scenarios of seeding, VE (as indicated in the subtitles), and NPIs (as indicated by the x-axis labels). All numbers are scaled for 1 million people. For the projected percentages for each variant and uncertainty bounds, see Table S1.

Tallies of model-projected infections (Fig 4B) and deaths (Fig S4) reveal four key determinants of future pandemic outcomes. First, simultaneous introduction of both the B.1.351 and P.1 variants would lead to larger increases of infections and mortality than sole introduction of either variant (Fig 4B and Fig S4B). This result indicates the importance of limiting the introduction of multiple VOCs. Second, maintaining very high vaccine efficacies against all variants is critical for mitigating the risk of a large resurgence in populations with relatively high vaccination coverage (e.g., compare the first three subplots in Fig 4B). Third, continued non-pharmaceutical preventive measures will reduce infection resurgence as municipalities reopen economies. For instance, even with high vaccine efficacy, a rapid, full reopening before a very large proportion of the population are fully vaccinated could lead to approximately three times as many infections as when reopening occurs more slowly (Fig 4A and Fig 4B first subplot). Four, reassuringly, while projected trends for COVID-19-related mortality are in general similar to those for infection (Fig S4A vs. Fig 4A), lower proportions of COVID-19-related deaths would occur due to the increased transmission of B.1.351 and/or P.1 (Fig S4B vs. Fig 4B; note the larger proportion of deaths due to B.1.1.7 than infections). This is due to the currently higher vaccination coverage among older adults who have been prioritized for vaccination in NYC similar to many other municipalities (see Table S4 for vaccination coverage by age group). This finding emphasizes the importance of vaccine effectiveness against VOCs and of prioritizing vulnerable populations for vaccination in order to prevent severe outcomes of COVID-19.

## DISCUSSION

Despite vaccine availability, the future trajectory of the COVID-19 pandemic remains uncertain due to the potential additional emergence and continued spread of multiple VOCs. To improve understanding of future pandemic dynamics, here we have developed and applied a comprehensive model-inference system to quantify key viral properties for three VOCs: B.1.1.7, B.1.351, and P.1. Our estimates for the B.1.1.7 variant are consistent with detailed epidemiological evidence (32.5 – 54.6% increase in transmissibility and minimal immune evasion estimated here vs. 30 – 50% increase in secondary attack rate based on contact tracing data^3, 4^ and little immune evasion based on laboratory and real-world vaccination data^5, 6, 15^). Our estimates of the level of immune evasion for the B.1.351 and P.1 variants are also consistent with antibody neutralization data suggesting both variants can evade prior immunity induced by infection and vaccination, though to a larger extent for the B.1.351 variant.^7, 9^ Here we provide joint quantification of immune escape and the change in transmissibility for both variants. Overall, the model-inference system estimates and model simulations suggest that both B.1.351 and P.1 are likely more competitive than the B.1.1.7 variant due to their greater propensity for immune escape. These estimates are consistent with observations from Qatar^15^ and Canada^16^ showing that the proportion of infections caused by B.1.351 and/or P.1 increased despite earlier introduction and dominance of B.1.1.7 in these populations. Therefore, in spite of the current widespread prevalence of B.1.1.7 in Europe and North America, importation of B.1.351 and/or P.1 to these regions could replace B.1.1.7 dominance and lead to a further increase of infections by either B.1.351, P.1 or both variants. Mass-vaccination with highly effective vaccines is thus crucial for mitigating the risk of future SARS-CoV-2 resurgence, particularly as economies re-open.

Our model-inference system estimates substantial immune escape for both P.1 and B.1.351. For P.1, the mean estimate is bounded by a very broad confidence interval; however, for B.1.351 the uncertainty is more constrained and indicates greater confidence that a substantial level of immune escape occurs. These latter findings are supported by vaccine clinical trial and real-world data. In particular, Shinde et al.^13^ found a similar likelihood of COVID-19, mostly due to B.1.351, among trial participants who were seropositive at enrollment (i.e., after the first wave) compared to those seronegative. Although potential differences in risk of exposure among the two comparison groups may have somewhat biased this finding, substantial repeat and breakthrough infection occurred due to B.1.351. Further, recent data from Qatar^15^ indicate that individuals receiving the full dosing regimen of the Pfizer BNT162b2 mRNA vaccine are at greater risk of breakthrough infection with the B.1.351 variant than B.1.1.7. Continued monitoring of the severity of both repeat and breakthrough infections is needed to more fully understand ongoing risks of VOCs to both health and economy. In addition, the Qatar study^15^ highlights the importance of full vaccination (i.e., administration of both doses for mRNA vaccines), as participants gained little protection against severe illness after the first vaccine dose (vs. ∼50% efficacy for B.1.1.7), even though full protection against severe illness was retained after two vaccine doses. It is thus likely critical that best vaccination protocols are followed in order to confer protection against variants with immune escape properties and that potential waning of vaccine-induced immunity over time is monitored.

In light of the spatial expansion of B.1.1.7, B.1.351, and P.1 and the potential emergence of other new variants, vaccination is paramount for controlling the COVID-19 pandemic. It is imperative that vaccine production, distribution, and administration proceed expeditiously, particularly in resource-limited settings. Without effective global control of the pandemic, continued transmission of SARS-CoV-2 will give rise to additional new variants and pose new threats to all. As vaccines are distributed and administered, a continuation of non-pharmaceutical preventive measures is needed to minimize infections among the unvaccinated. As shown in our simulations, despite the relatively high vaccine coverage obtained to date in a place like NYC, COVID-19 infections could resurge if such locations lift preventive measures prematurely.

Due to a lack of sub-regional data, we used aggregated country-level data to estimate the properties for the three VOCs. Our model-inference system also did not account for differences of disease severity and infection-detection rate among age groups, which may vary substantially.^17^ This model simplification may introduce uncertainty and bias to our estimates, particularly for Brazil where country-level data may mask more intense transmission in subpopulations and may have led to underestimation of the transmissibility of P.1. Nevertheless, validation using independent data, including for Brazil, indicates that the model-inference system is able to closely capture pandemic dynamics and accurately estimate cumulative infection rates (Fig 2). Further, because the model-inference system simultaneously accounts for population susceptibility, disease seasonality, NPIs, and vaccination, it is able to specifically estimate changes related to a given new variant and closely matches available epidemiological data (e.g., for B.1.1.7). The model simulations using these estimated characteristics further illustrate the relative competitiveness of the three VOCs and delineate key determinants of future infection outcomes. Overall, our findings point to the importance of preventing the spread of the B.1.351 and P.1 variants, in addition to B.1.1.7, via continued NPIs, prompt mass-vaccination of all populations, continued monitoring of vaccine efficacy, and potentially updating vaccine formulations to ensure high efficacy.

## METHODS

### Data sources and processing

The model-inference system uses reported COVID-19 case and mortality data to capture transmission dynamics, weather data to estimate disease seasonality, mobility data to represent concurrent NPIs, and vaccination data to account for changes in population susceptibility due to vaccination, for each of the three countries (i.e., the UK, South Africa, and Brazil). Country-level daily COVID-19 case and mortality data came from COVID-19 Data Repository of the Center for Systems Science and Engineering (CSSE) at Johns Hopkins University;^18, 19^ we aggregated the data to weekly intervals until the week of 4/11/2021 but excluded initial weeks with low case rates (<2 per million population). Hourly surface station temperature and relative humidity came from the Integrated Surface Dataset (ISD) maintained by the National Oceanic and Atmospheric Administration (NOAA) and are accessible using the “stationaRy” R package.^20, 21^ We computed specific humidity using temperature and relative humidity per the Clausius-Clapeyron equation.^22^ We then aggregated these data for all weather stations in each country with measurements since 2000 and calculated the average for each week of the year during 2000-2020. To compute the seasonal trend, we used a method developed by Yuan et al.^23, 24^ which estimates the relative reproduction number based on temperature and specific humidity (see details in Supplemental Information). Daily mobility data were derived from Google Community Mobility Reports;^25^ we aggregated all business-related categories (i.e., retail and recreational, transit stations, and workplaces) in all locations in each country to weekly intervals. Daily vaccination data (for 1^st^ and 2^nd^ dose if applied) for the UK were sourced from Public Health England;^26^ and data for South Africa and Brazil were obtained from Our World in Data.^27, 28^

### Model-inference system

Contact tracing data capturing chains of transmission can be used to compute the secondary attack rate and quantify changes in transmissibility due to a given new variant. Surveillance data and laboratory viral characterization can be used to document and quantify levels of immune evasion. Yet such detailed data are often not available, particularly for resource-limited settings. Given these circumstances, mathematical modeling that assimilates epidemic surveillance data provides an attractive alternate means for estimating key epidemiological properties of novel variants, including the transmission rate. However, joint estimation of the transmission rate and population susceptibility, which is related to immune evasion, is challenging, as both quantities can change for a new variant. This problem arises mathematically because the product of these two quantities, rather than either individually, determines disease incidence – i.e., at any point in time, given the incidence, transmissibility and susceptibility are not individually identifiable. Nevertheless, we posit that transmissibility and susceptibility affect epidemic dynamics differentially *over time*, and, as such, data at multiple time points can enable joint estimation. We thus design a model-inference system to estimate the most plausible joint changes in these two quantities using commonly available incidence and mortality time series.

The model-inference system is comprised of an epidemic model for simulating the transmission dynamics of SARS-CoV-2 and a statistical inference method for estimating the model state variables and parameters. The epidemic model is a susceptible-exposed-infectious-recovered-susceptible (SEIRS) construct that further accounts for two-dose vaccination. In addition, to include the effects of public health interventions and disease seasonality, it further adjusts the transmission rate each week using mobility data and the estimated seasonal trend based on climate conditions (see Eqn S3 in the Supplemental Information). The system then combines the model-simulated (prior) estimates and observed case and mortality data to compute posterior estimates using the ensemble Kalman adjustment filter (EAKF).^29^ We also apply a technique termed space re-probing^30^ that accommodates possible large changes mid-pandemic to transmissibility and population susceptibility. Further, due to the challenge identifying these two quantities individually, we ran the model-inference system, repeatedly and in turn, in order to test 14 major combinations of changes in transmissibility and susceptibility (see details in Supplemental Information). Briefly, depending on the hypothesized change, we restricted the EAKF update to a given related set of parameters or variables. For instance, for the hypothesis that the new variant changes the transmissibility but does not evade immunity, the system only allows major adjustment to the transmission rate and the infectious period; for the hypothesis that the new variant induces both changes, the system allows major adjustment to the transmission rate, the infectious period, and population susceptibility. The system then selects the run with the best performance based on the accuracy of model-fit, one-step ahead prediction, and magnitude of changes to key state variables to identify the most plausible combination of changes in transmissibility and level of immune evasion (see Supplemental Information). To approximate the distribution of the system (including all model state variables and parameters), we employed an ensemble of model replica (n = 500 here) and updated the ensemble posterior each week using the EAKF. In addition, to account for model stochasticity, we repeated each model-inference simulation 100 times for each dataset, each with initial parameters and variables randomly drawn from prior distributions (Table S2). Consequently, model estimates are aggregated from 50,000 model runs in total.

### Estimation of variant-specific changes in transmissibility and level of immune evasion

The model decouples the impact of concurrent NPIs and disease seasonality from the transmission rate and infectious period (Eqn S3); as such, estimates for the latter two parameters are variant-specific. We thus compute transmissibility as the product of the transmission rate and infectious period. To reduce uncertainty, we average transmissibility estimates over the first pandemic wave (excluding the first three weeks when model estimates are less accurate) for the wild-type SARS-CoV-2 virus; similarly, we average the transmissibility estimates over the period when the new variant is dominant. We identify this latter time period based on the transmissibility estimates: 1) For the UK (B.1.1.7) and South Africa (B.1.351), the estimated transmissibility increased and plateaued within 10 weeks (see Fig 3); we thus used the period from the week with the maximal transmissibility during the 10 weeks following its initial increase to the end of our study period (i.e., the week of 4/11/2021). However, for the UK, we excluded the 3^rd^ lockdown period when estimated transmissibility is lower, potentially due to better awareness of B.1.1.7 and preventive measures taken at the time not fully captured by the model. Of note, contact tracing data also indicate a lower increase of the secondary attack rate around that time: 25-40% during 11/30/20 – 1/10/21 (among 1,364,301 cases for this expanded analysis)^4^ vs. 30-50% during 11/30 – 12/20/20 (among 386,805 cases).^3^ 2) For Brazil (P.1), the estimated transmissibility increased more gradually (see Fig 3), we thus instead used either the weeks identified per 1) or the last 8 weeks of our study period, whichever with a longer time period, to ensure the robustness of estimation. We then compute the average change in transmissibility due to a new variant as the ratio of the two averaged estimates (i.e., after: before the rise of the new variant).

To quantify immune evasion, we record all time points inducing major EAKF adjustments to posterior estimates of susceptibility, compute the change in immunity as ΔImm = *S_t+1_* – *S_t_* + *i_t_* (with *S_t_* as the susceptibility at time-*t* and *i_t_* as the new infections occurring at time-*t*), and sum over all ΔImm estimates to compute the total change in immunity due to the new variant. We then compute the level of immune evasion as the ratio of the total change in immunity during the second wave to the model-estimated population immunity at the end of the first wave. This ratio provides an estimate of the fraction of individuals previously infected who are susceptible to re-infection with the new variant.

For both quantities, we report the mean and 95% CI based on the mean estimates from 100 repeated model-inference runs.

### Model validation using model-generated synthetic data

To test the accuracy of the model-inference system, we generated 10 synthetic datasets using a two-variant SEIRS model (see Eqn. S4) and different scenarios of changing transmissibility and immune evasion (Table S3). In each scenario, a new variant was introduced at week 21 of the simulation. We then combined the incidence and mortality due to both variants and added noise drawn from a Poisson distribution to represent observational error. We then applied the model-inference system to estimate the model variables and parameters for each synthetic dataset, per the procedure described above for real data. For comparison with model-inference system estimates, we computed the true values of population susceptibility and transmissibility over time as the weighted average of the two variants based on the relative prevalence at each time point (i.e., each week).

### Model validation using independent data

To compare model estimates with independent observations not assimilated into the model-inference system, we identified four relevant datasets: 1) the REACT study, which measures the prevalence of SARS-CoV-2 using PCR-testing of volunteers from the general public living in the UK. At the time of this study, the REACT study has conducted 10 rounds of testing during 5/1/2020 – 3/30/2021 (n = 1,572,951 tests in total)^10, 11^; Fig 2B plots our estimates of prevalence of SARS-CoV-2 each week, overlaying all 10 measures from the REACT study for corresponding time periods; 2) a serosurvey of workers in Cape Town, South Africa, conducted during 8/17 – 9/4/2020 (n = 405 participants);^12^ 3) serology tests among participants enrolled during 8/17 – 11/25/2020 in the Novavax NVX-CoV2373 vaccine phase 2a-b trial in South Africa (n=1324).^13^ Given this long enrollment period, we used the centered 2-week window (9/29 – 10/13/20) to match with our model estimates. and 4) two nationwide random household serosurveys conducted in Brazil during 5/14 – 5/21/2020 (n = 25,025 participants) and 6/4 – 6/7/2020 (n = 31,165 participants).^14^ To account for the delay in antibody generation, we shifted the timing of each serosurvey 14 days when comparing to model-inference system estimates of cumulative infection rates in Fig 2 D and E.

### Model simulations testing the relative competitiveness of VOCs and projecting future transmission dynamics

Here we modified a multi-variant model previously developed for influenza virus^31^ to include age structure and interactions (Eqn. S5). The multi-variant model accounts for: 1) competition between each pair of SARS-CoV-2 variants (e.g. wild-type and B.1.1.7) via cross-protective immunity; 2) variant-specific transmissibility and population susceptibility, based on estimates derived in this study; 3) variant-specific vaccine efficacy under different scenarios (see Table S5); 4) age-specific differences in vaccination coverage at the start of simulation and vaccination uptake rates for the simulation period (see Table S4); 5) seasonality; and 6) changes in NPIs under different scenarios (see Table S5). We used data from NYC for baseline vaccination coverage^32^ and initial prevalence of different variants,^33^ as well as key model estimates (e.g., transmission rates and infection fatality risk by age group; see Table S5)^17, 34, 35^. As in previous work,^17, 34, 35^ we included 8 age-groups (i.e. <1, 1-4, 5-14, 15-24, 25-44, 45-64, 65-74, and 75+ year-olds) in the model to account for age-specific differences. To focus on the three VOCs, we only included the B.1.1.7, B.1.351, and P.1 variants and attributed all other variants as “wild-type” virus, even though at the start of the simulations, the B.1.526 variant made up approximately one third of sequenced infections (N.b., the B.1.526 variant likely emerged locally in NYC; we estimated a ∼20% increase in transmissibility and nominal immune evasion for this variant; based on these estimates the impact of this variant is expected to be relatively minor). We did not account for potential differences in infection fatality risk by variant, as such information is not available; therefore, the simulated mortality under different scenarios only reflect the relative infection rate by age group, for which we apply age-specific infection-fatality risk (see Table S5). In addition, due to uncertainty vis-à-vis the severity and infection fatality risk among breakthrough infections (i.e., those who have been vaccinated), we only show mortality-related simulations for the “Same VE” scenario which assumes no reduction in VE.

### Data Availability

All data used in this study are publicly available as described in the “Data sources and processing” section.

### Code availability

All source code and data necessary for the replication of our results and figures will be made publicly available.

## Data Availability

All data used in this study are publicly available as described in the Data sources and processing section.

## Acknowledgements

This study was supported by the National Institute of Allergy and Infectious Diseases (AI145883), the National Science Foundation Rapid Response Research Program (RAPID; DMS2027369) and a gift from the Morris-Singer Foundation.

## Author contributions

WY designed the study (main), conducted the model analyses, interpreted results, and wrote the first draft. JS designed the study (supporting), interpreted results, and critically revised the manuscript.

## Supplemental Information

### 1. Estimating seasonal trends

Many respiratory infections tend to occur seasonally and are predominantly prevalent during certain months of the year (e.g., cold months in temperate climates).^1^ This seasonal pattern has been documented for influenza viruses,^2^ respiratory syncytial viruses,^3^ and endemic human coronaviruses.^4^ In addition, studies have showed that this seasonality may be associated with climate conditions – particularly, temperature and humidity – as they may modulate the survival and transmission of respiratory viruses.^5–8^ For the SARS-CoV-2 virus, our work has also shown that a winter-time seasonality exists, similar to endemic human coronaviruses in New York City (NYC), and that models accounting for this seasonality enable more accurate projection of COVID-19 pandemic dynamics than those do not.^9, 10^ However, to date, no mechanistic models exist that quantify the response of the SARS-CoV-2 virus to temperature and humidity and in turn the seasonality of COVID-19. In addition, seasonal trends may differ by climate. For instance, epidemics of influenza can occur any time of the year in subtropical and tropical climates; it is thus more difficult to characterize the seasonality of respiratory infections in these climates. To address these challenges, we recently developed a flexible climate-forced model of epidemic dynamics for subtropical and tropical climates; results with this model also describe the response to temperature and humidity conditions common in temperate climates.^11, 12^ Thus, to account for the potentially diverse seasonal trends of COVID-19 in the UK (temperate climates), South Africa (mostly temperate climates), and Brazil (mostly tropical climates), we applied this climate-forcing to temperature and humidity data for each country and computed the relative seasonal trend for each country.

Specifically, the climate-forcing takes the following form:

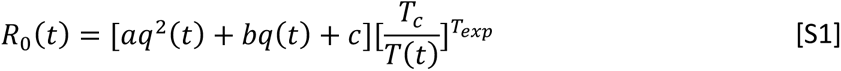

where *R_0_*(*t*) is the basic reproduction number at time *t*; *q(t)* is specific humidity (i.e. a measure of absolute humidity) at time *t*; and *T(t)* is temperature at time *t*. In essence, the forcing function assumes that specific humidity has a bimodal effect on *R_0_*, with both low and high humidity conditions favoring transmission; in addition, this effect is moderated by temperature, where low temperatures promote transmission and temperatures above a certain threshold (i.e., *T_c_* in Eqn. S1) limit transmission. Further, to link the coefficients *a*, *b*, and *c* to humidity *q* and *R_0_*, Yuan et al.^11, 12^ reparametrized the forcing function by solving the parabola with a nadir at (*q_mid_*, *R_0max_ -R_0diff_*) and maxima at both (*q_min_*, *R_0max_*) and (*q_max_*, *R_0max_*), such that:

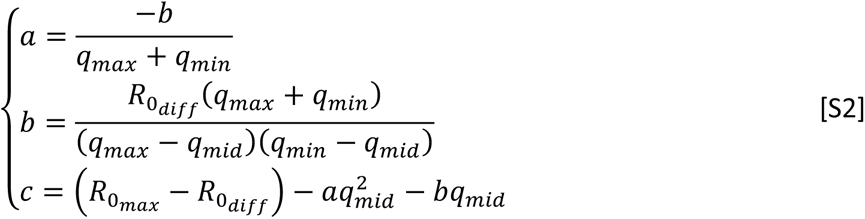

Yuan et al.^11, 12^ estimated the parameters *R_0max_* (i.e., the maximum *R_0_*), *R_0diff_* (i.e., the difference between the maximum and minimum *R_0_*), *q_min_*, *q_mid_*, and *q_max_* (i.e., the minimum, median, and maximum specific humidity for the response), *T_c_* (the threshold temperature) and *T_exp_* (the exponent in Eqn S1) for influenza in Hong Kong, a subtropical city, based on long-term epidemic data collected therein during 1998 - 2018. Here we use their mean estimates for these parameters and temperature and humidity data for each country (see main text and Fig S5) to compute the seasonal trend for each country using Eqns. S1-2. However, as these parameters were estimated for influenza, the outputs do not represent the actual *R_0_* for the SARS-CoV-2 virus. Thus, we instead compute the *relative* seasonal trend, by scaling the weekly country output from Eqn S1 by the country mean output, such that this scaled output provides a *relative*, seasonality-related transmissibility for each week of the year (see results in Fig S2). These relative estimates also decouple the seasonality-related and variant-specific transmissibility (assuming no interaction; see below).

### 2. Model-inference system

The model-inference system developed for this study consists of an SEIRSV model to simulate the transmission dynamics of SARS-CoV-2 and the ensemble Kalman adjustment filter (EAKF)^13^ to estimate the model state variables and parameters, based on case and mortality data. Here we describe the model and the filtering method in detail.

#### 2.1. Epidemic model

The SEIRSV (susceptible-exposed-infectious-recovered-susceptible-vaccination) model uses the following set of equations to simulate the transition of sub-populations between different disease stages, while accounting for disease seasonality, concurrent non-pharmaceutical interventions (NPIs), and vaccination:

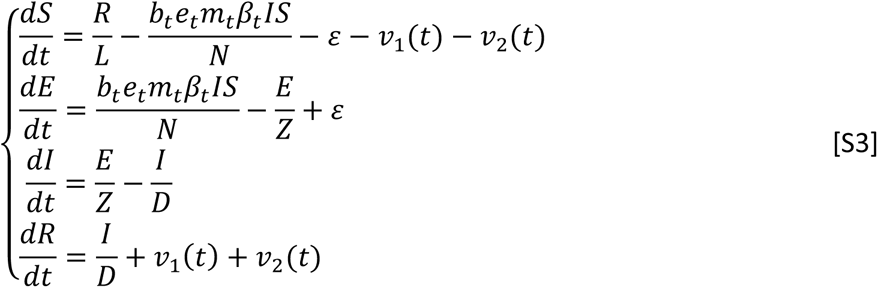

where *S*, *E*, *I*, *R* are the number of susceptible, exposed (but not yet infectious), infectious, and recovered/deceased individuals, respectively; *N* is the population size. The parameter *ε* represents travel-related importation of infections (nominally set to 1 per 20 days per 1 million population, unless specified otherwise). To account for local seasonality, *b_t_*, the estimated relative seasonal trend for each country (see Section 1 above and Fig S2) is used to adjust the relative transmission rate at time *t*. To account for concurrent NPIs, the term *m_t_*, the relative population mobility at time *t* (in this study, we use data from Google Community Mobility Reports;^14^ see main text and Fig S2), is used to adjust the transmission rate. In addition, as the effectiveness of NPIs is unknown and variable, the model further includes a parameter, *e_t_*, to scale NPI effectiveness at time *t*. For virus-specific characterization, N_K_ is the variant-specific transmission rate at time *t*, *Z* is the latency period, *D* is the infectious period, and *L* is the immunity period. Note that the parameters *e_t_*, N_K_, *Z*, *D*, and *L* are estimated by the model-inference system as described below.

To incorporate changes in population susceptibility due to vaccination, the model accounts for two-dose vaccination via *v_1_*(*t*) and *v_2_*(*t*). Specifically, *v_1_*(*t*) is the number of individuals successfully immunized after the first dose of the vaccine and is computed using vaccination data and vaccine efficacy for 1^st^ dose (see detailed settings in Table S2). Similarly, *v_2_*(*t*) is the additional number of individuals successfully immunized after the second vaccine dose (excluding those successfully immunized after the first dose).

#### 2.2. Observation model

We compute the model-simulated number of cases and deaths for each week using the model-simulated infection rate, as done in Yang et al.^10^ Specifically, we include 1) a time-lag from infectiousness to detection (i.e., an infection being diagnosed as a case) – drawn from a gamma distribution with a mean of *T_m_* and standard deviation (SD) of *T_sd_* days – to account for delays in diagnosis and detection; 2) an infection-detection rate (*r*), i.e. the fraction of infections (including subclinical or asymptomatic infections) reported as cases, to account for under-detection; 3) a time-lag from infectiousness to death, drawn from a gamma distribution with a mean of 14 days and a standard deviation of 10 days, empirically based on mortality data; and 4) an infection-fatality risk (IFR), i.e., the fraction of infections that die from COVID-19. To compute the model-simulated number of new cases per week, we multiply the model-simulated number of new infections per day by the infection-detection rate, and further distribute these simulated cases in time per the distribution of time-from-infectiousness-to-detection. We then aggregate the daily lagged, simulated cases to weekly totals for model inference (see below). Similarly, to compute the model-simulated deaths per week and account for delays in time to death, we multiply the simulated-infections by the IFR and then distribute these simulated deaths in time per the distribution of time-from-infectiousness-to-death, and aggregate these daily numbers to weekly totals. For each week, the infection detection rate (*r*), the mean (*T_m_*) and standard deviation (*T_sd_*) of time-from-infectiousness-to-detection, and the IFR are estimated based on weekly case and mortality data, along with other model parameters.

#### 2.3. Inference using the EAKF

At the end of each week, the inference system uses the EAKF to update the state variables and parameters based on model-generated prior estimates and case and mortality data. Briefly, the EAKF uses an ensemble of model realizations (*n*=500 here), each with initial parameters and variables randomly drawn from a prior range (see Table S2). After model initialization, the system integrates the model ensemble forward in time for a week (per Eqn S3) *stochastically* to compute the prior distribution for each model state variable or parameter, as well as the model-simulated number of cases and deaths for that week as described in Section 2.2. The system then combines the prior estimates with the observed case and death data for the same week to compute the posterior per Bayes’ theorem.^13^ During this filtering process, the system updates the posterior distribution of all model parameters and variables for each week.^13^ As such, it is able to capture the time-varying changes in transmission dynamics including the variant-specific transmission rate (N_K_) and infectious period (*D*) – the two parameters that we use to compute variant-specific transmissibility over time.

However, unlike previous studies using similar model-inference approaches, here we further modify the EAKF filtering process to test different potential combinations of changes in transmissibility and immune escape. To enable this exploration of systemic changes (e.g. due to the emergence of a new variant), we randomly replace a small fraction of ensemble members (3-10%) using values randomly drawn from specified ranges. This technique, termed space reprobing (SR), was developed in order to explore state space without corrupting performance of the filter.^15^ Specifically for this application, we apply SR to a given related set of parameters/variables and restrict the EAKF update of non-related parameters/variables, for 14 different hypothesized behaviors. These hypothesized changes are as follows:

1. Hypothesis 1 (minor changes in transmissibility, no immune escape): Large updates are only allowed for the two transmissibility-related parameters N_K_ and *D*; to explore the changes, the system applies SR to these two parameters using values drawn from prior ranges 10-20% higher than the initial priors.
2. Hypothesis 2 (major changes in transmissibility only, no immune escape): Similar to 1); but to explore the changes, the system applies SR to N_K_ and *D* using values drawn from prior ranges 30-40% higher than the initial priors.
3. Hypothesis 3 (minor immune escape only, no changes to transmissibility): Large updates are only allowed for *S*, the population susceptibility, up to a total loss of 50% of the prior immunity.
4. Hypothesis 4 (major immune escape only, no changes to transmissibility): Large updates are only allowed for *S*, the population susceptibility, up to a total loss of 95% of the prior immunity.
5. Hypothesis 5 (minor changes in transmissibility + minor immune escape): combining 1) and 3) above.
6. Hypothesis 6 (major changes in transmissibility + major immune escape): combining 2) and 4) above.
7. Hypothesis 7 (minor changes in transmissibility + major immune escape): combining 1) and 4) above.
8. Hypothesis 8 (major changes in transmissibility + minor immune escape): combining 2) and 3) above.
9. Hypothesis 9 (changes in both transmissibility and immune escape, no restriction on magnitude of change): Large updates are allowed for N_K_ and *D* as well as *S*. To explore the changes, initial SR uses values drawn from prior ranges 10-20% higher than the initial priors, and values up to 30-40% higher than the initial priors if the inference system detects the prior continues to underestimate the observed cases and deaths with the 10-20% initial increase in SR values. In addition, SR allows updates of *S* up to 95% of the prior immunity. To account for slower changes in overall population immunity (i.e., in the entire country) as the new variant gradually spreads to different sub-regions across a large geographic space, such as in Brazil, we also explore the fitting using the following five additional settings:
10. Hypothesis 10 (immune escape only and changes to overall population immunity occur slowly over time): Large updates are only allowed for *S*, up to a total loss of 95% of the prior immunity; however, SR is applied to a smaller fraction of ensemble members than in 1)-9) such that changes in *S* occur gradually.
11. Hypothesis 11 (minor changes in transmissibility + minor immune escape; both occur slowly over time): Large updates are allowed for N_K_ and *D* as well as *S*. Adjustment to *S* is allowed as in 10) but up to only 50% of prior immunity. In addition, for transmissibility, the system applies SR to N_K_ and *D* using values drawn from prior ranges 10-20% higher than the initial priors.
12. Hypothesis 12 (major changes in transmissibility + minor immune escape; both occur slowly over time): Similar to 11); however, for N_K_ and *D*, initial SR uses values drawn from prior ranges 10-20% higher than the initial priors, and values up to 30-40% higher than the initial priors if the inference system detects the prior continues to underestimate the observed cases and deaths with the 10-20% initial increase in SR values.
13. Hypothesis 13 (minor changes in transmissibility + major immune escape; both occur slowly over time): Similar to the settings specified in 11) but adjustment to *S* is allowed up to 95% of prior immunity.
14. Hypothesis 14 (major changes in transmissibility + major immune escape; both occur slowly over time): Similar to settings specified in 12) but adjustment to *S* is allowed up to 95% of the prior immunity.

We carry out the model-inference process for each of the 14 settings described above and for each country dataset. We then select the most plausible hypothesis for each country based on the following criteria: 1) model fitting to case and mortality data, as indicated by the relative root-mean-squared-error (RRMSE) between the *posterior* estimates for the corresponding variable (i.e. case rate or mortality rate) and data; 2) the accuracy of one-step ahead prediction, as indicated by the RRMSE between the *prior* estimates for the corresponding variable (i.e. case rate or mortality rate) and data; 3) the level of adjustment needed for two key variables, i.e., infection rate and case rate, as indicated by the RRMSE between the *prior* and *posterior* estimates for each variable; 4) a penalty on the number of variables needing SR adjustment; and 5) a penalty on the frequency of SR adjustment. We combine all these metrics by weighting them heuristically using the following set of weights: 0.27 for the two metrics in 1); 0.13 for the two metrics in 2), 0.03 for the two metrics in 3), and 0.07 for both 4) and 5). We also tested other sets of weights and found that higher weights should be given to 1) and 2) based on results from the synthetic testing where the ‘true’ values of the state variables and parameters are known; however, in general, the final results are similar if there are minor changes to these weights.

To account for model stochasticity, we repeat each model-inference 100 times for each dataset, each with initial parameters and variables randomly drawn from the prior distributions (Table S2). Each model-inference tests the 14 hypotheses described above, selects the one with the best performance (i.e. minimizing the combined metric described above), and outputs the estimates of the best-performing run. That is, the model estimates reported in the main text are aggregated from 100 best-performing model runs (each with 500 ensemble members and totaling 50,000 individual model realizations).

### 3. Model validation using model-generated synthetic data

To test the accuracy of our model-inference system, we generate 10 synthetic datasets using a separate multi-variant SEIRS model, similar to models developed in Yang et al.^16^ and Gog & Grenfell.^17^ Within this model, variants can interact via cross-immunity, which protects a portion of individuals with prior infection (i.e. polarized immunity) by reducing transmission.

Specifically, the model takes the following form:

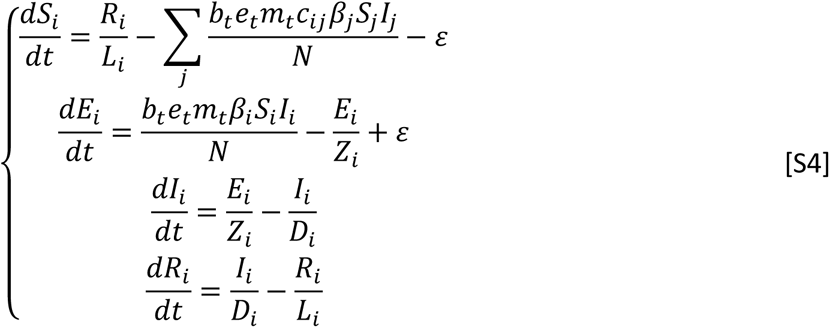

where *N* is the population size; *S_i_*, *E_i_*, *I_i_*, and *R_i_*, are, respectively, the numbers of susceptible, exposed-but-not-yet infectious, infectious, and recovered individuals, with respect to variant-*i* (here, the wild-type SARS-CoV-2 virus or a new variant); N_=_, *D_i_*, and *L_i_* are, respectively, the transmission rate, mean infectious period, and mean immunity period, for variant-*i*; and *c_ij_* measures the strength of cross-immunity to variant-*i* conferred by infection with variant-*j* (e.g., close to 0 if it is weak and *c_ii_*=1 for infection by the same variant). The parameter *ε* represents travel-related importation of infections; to generate the synthetic data (i.e., “truths”), we set *ε* to 1 per week for the first 5 weeks and 1 every 3 days for the rest of the first wave (here weeks 1-20); for the second wave, as transmission has been established locally, we set *ε* to 0 for simplicity. The terms *b_t_*, *m_t_*, and *e_t_* are the same as in Eqn S3 and account for seasonality and NPIs over time. For simplicity, we omit birth, death, and vaccination.

To generate the synthetic data (i.e., “truths”), we seed the Eq. S4 model with 2 infections of wild-type virus at the start of each simulation and 50 infections of a new variant at the start of Week 21, for *N* = 1 million people; we run the model stochastically with a daily time-step from the week starting 3/1/2020 to the week starting 2/21/2021 (i.e. 52 weeks in total) using the parameters listed in Table S3. To compute the weekly number of cases and deaths, we use the same procedure as described in Section 2.2 above for each variant. We then combine the case/mortality estimates for both variants, add random noises drawn from a Poisson distribution to mimic observational error. The final noise-added weekly case and mortality time series are then used as synthetic data for testing the model-inference system (described in Section 2 above). To compare the posterior estimates of key parameters and variables (e.g. transmissibility and population susceptibility) from the model-inference system, we compute the true values of population susceptibility and transmissibility over time as the weighted average of the two variants based on the relative prevalence during each week. Fig 1 and Fig S1 show the 10 model-generated truths including cases, deaths, and the computed “true” values of population susceptibility and transmissibility for each week of the simulation.

### 4. Multi-variant, age-structured model for simulation to test the relative competitiveness of VOCs and project future SARS-CoV-2 dynamics

We modify the multi-variant model in Eqn S4 to further include age structure and vaccination. The inclusion of age structure here allows incorporation of age-specific parameters (e.g., transmission rate and infection-fatality risk) as well as age-specific vaccination coverage and rates. Specifically, this multi-variant, age-structured SEIRSV model takes the following form:

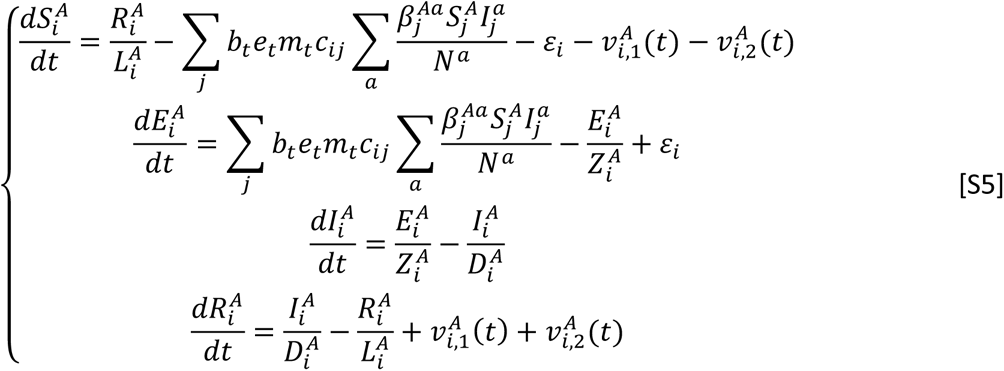

Model parameters in Eqn S5 are similar to those in Eqn S4, expect for those related to age, which are indicated by the superscripts. The vaccination model component is also similar to Eqn S3, but with age-stratification. However, the terms 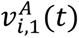 and 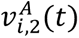 are variant-specific, as indicated by the additional subscript *i*; that is, they additionally account for the reduction in vaccine efficacy against the new variants, based on scenario assumptions specified in Table S5.

As an example, we simulate the transmission dynamics under different scenarios of variant prevalence, vaccine efficacy, and NPIs for a city like NYC, from the week of 4/25/2021 to the week of 8/22/2021 (i.e., approximately May - August 2021). We use data or estimates available for NYC to initialize the parameters and state variables needed for model simulations. In addition, we use our model-inference estimates for the VOCs for related parameters and variables. Specifically, as in previous work,^9, 10, 18^ we include 8 age-groups (i.e. <1, 1-4, 5-14, 15-24, 25-44, 45-64, 65-74, and 75+ year-olds) to account for age-specific differences. To focus on the three VOCs, here we only include the B.1.1.7, B.1.351, and P.1 variants and attribute the rest as “wildtype” virus. NYC data on variant prevalence among tested infections during the weeks of March – April 2021 are used to inform the initial range of seeding for each variant (Table S5). Initial conditions for the state variables (e.g., susceptibility and SARS-CoV-2 prevalence) for each age group are taken from estimates^9, 10, 18^ made for the week of 4/18/2021, using detailed data (including case, mortality, COVID-19-related emergency visit, mobility, and vaccination) during 3/1/2020 – 4/24/2021. For each age group, to compute the initial variant-specific population susceptibility (*S_i_*) at the start of a simulation, we move the estimated proportion with immune escape for variant *i* among those who have had prior infection with the wild-type virus but have not been vaccinated back to the susceptible compartment. The number of people losing vaccine-induced immunity is computed based on scenario assumptions determining the reduction in vaccine efficacy (see scenarios in Table S5).

The cross-immunity settings, i.e., values of *c_ij_*’s in Eqn S5, come from our posterior model-inference estimates of immune escape and are used for all age groups. To reduce uncertainty, here we use the 80% CI estimates (see Table S5). For instance, as our model-inference estimate of immune escape for B.1.351 has an 80% CI of 40.1 – 82.8%, we set *c*(B.1.351←wildtype), the cross-immune protection against B.1.351 conferred by prior infection of the wildtype virus relative to variant-specific immunity, to values drawn from a uniform distribution ranging from 0.172 to 0.599 (i.e. cross-immunity is set to the complement of estimated immune escape). We set all *c*(wildtype←new variant) to 1 – that is, we assume full cross-immune protection against the wild-type virus conferred by infection due to any VOC.

Similarly, the variant-specific transmission rates, i.e. N_=_’s, come from our posterior model-inference estimates of the relative transmissibility for each variant. For instance, as our model-inference estimate of transmissibility for B.1.351 is 18.5 – 45.7% (80% CI) higher than that of the wildtype virus, we set N_B.1.351_ to 1.185 – 1.457 times of the estimate for N_wildtype_. The same scaling is applied to all age groups.

Due to a lack of information, we do not account for potential differences in infection fatality risk by variant; therefore, the simulated mortality under different scenarios only reflect the relative infection rate by age group, for which we apply age-specific infection-fatality risk (see Table S5). In addition, due to the uncertainty of the infection fatality risk among breakthrough infections (i.e., those who have been vaccinated), we only show mortality-related simulations for the “Same VE” scenario which assumes no reduction in VE.

We run the model for each scenario 1000 times stochastically, with the parameters and initial conditions randomly drawn from uniform distributions with ranges specified in Table S5. Results are summarized from the 1000 model runs for each scenario.

---

**Fig S1.**
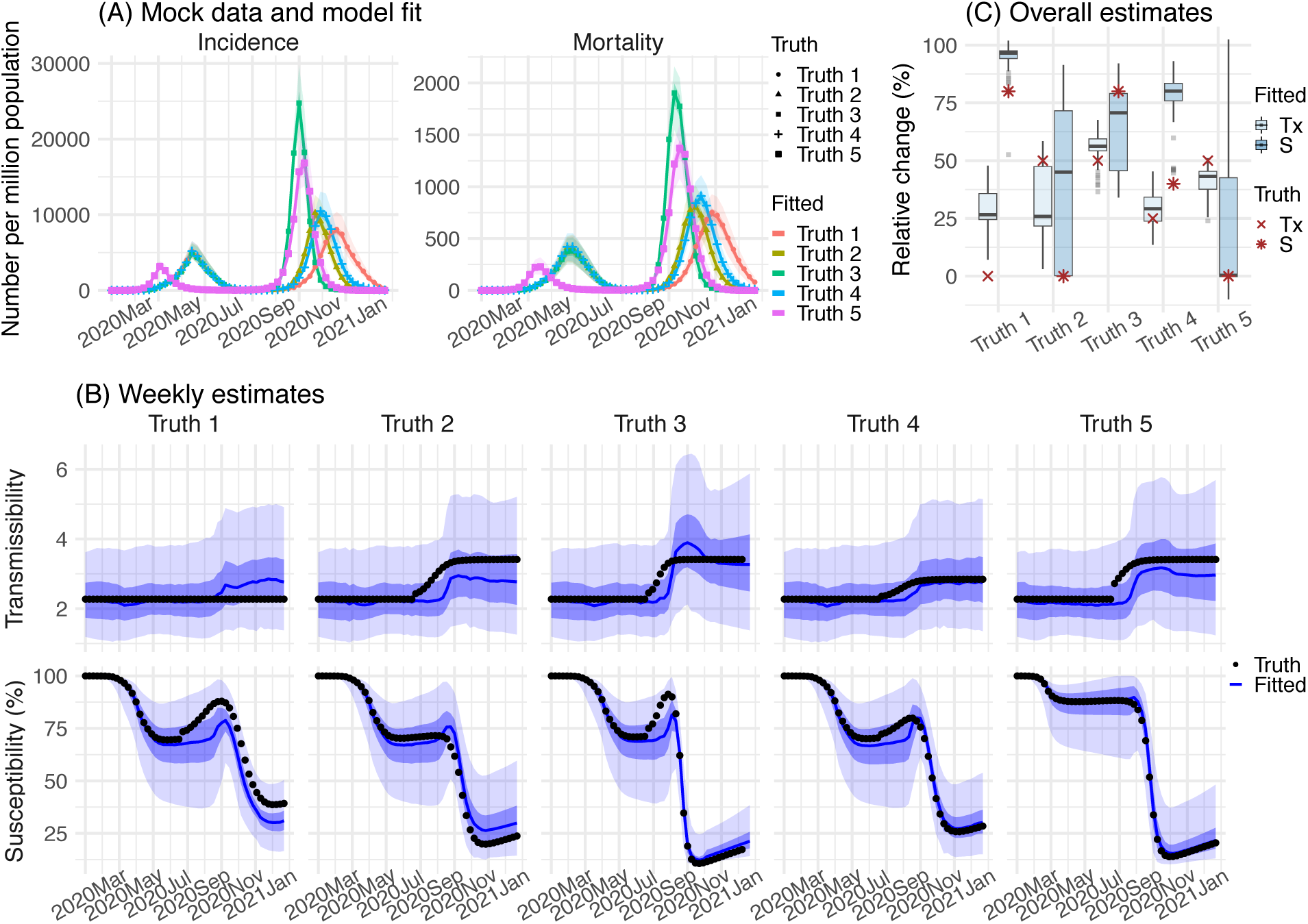
Model validation using model-generated synthetic data with an infection-detection rate set to 10%. (A) 5 sets of synthetic data (dots) and model-fits to each dataset; lines show mean estimates and surrounding areas show 50% (dark) and 95% (light) CrIs. (B) weekly model estimated transmissibility and population susceptibility; lines show mean estimates and surrounding areas show 50% (dark) and 95% (light) CrIs, compared to the true values (dots). (C) overall estimates of the change in transmissibility and immune evasion (boxes and whiskers show model estimated median, interquartile range, and 95% CI from 100 model-inference simulations) compared to the true values (dots).

**Fig S2.**
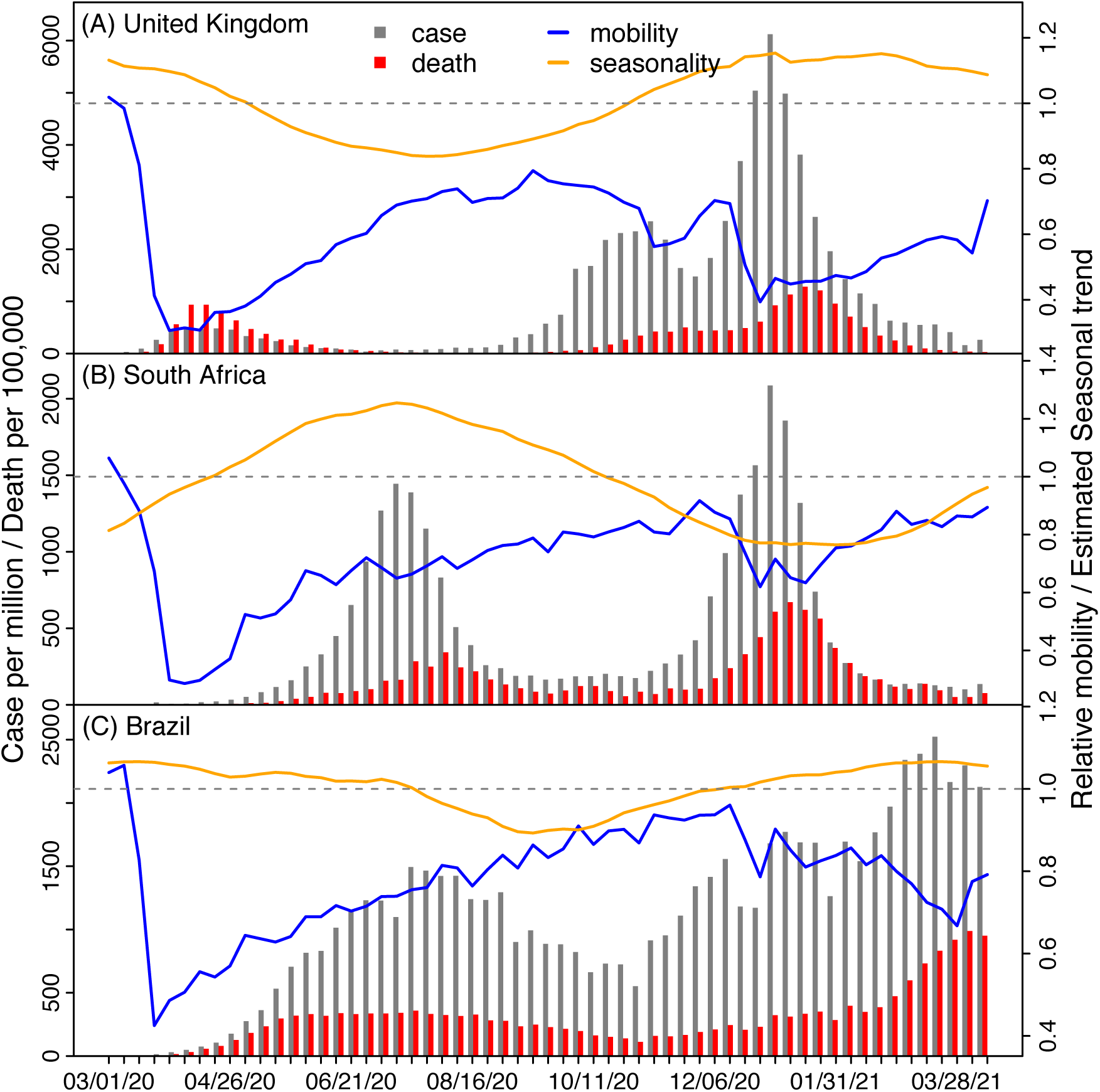
Pandemic dynamics, mobility, and estimated seasonal trends in the three countries.

**Fig S3.**
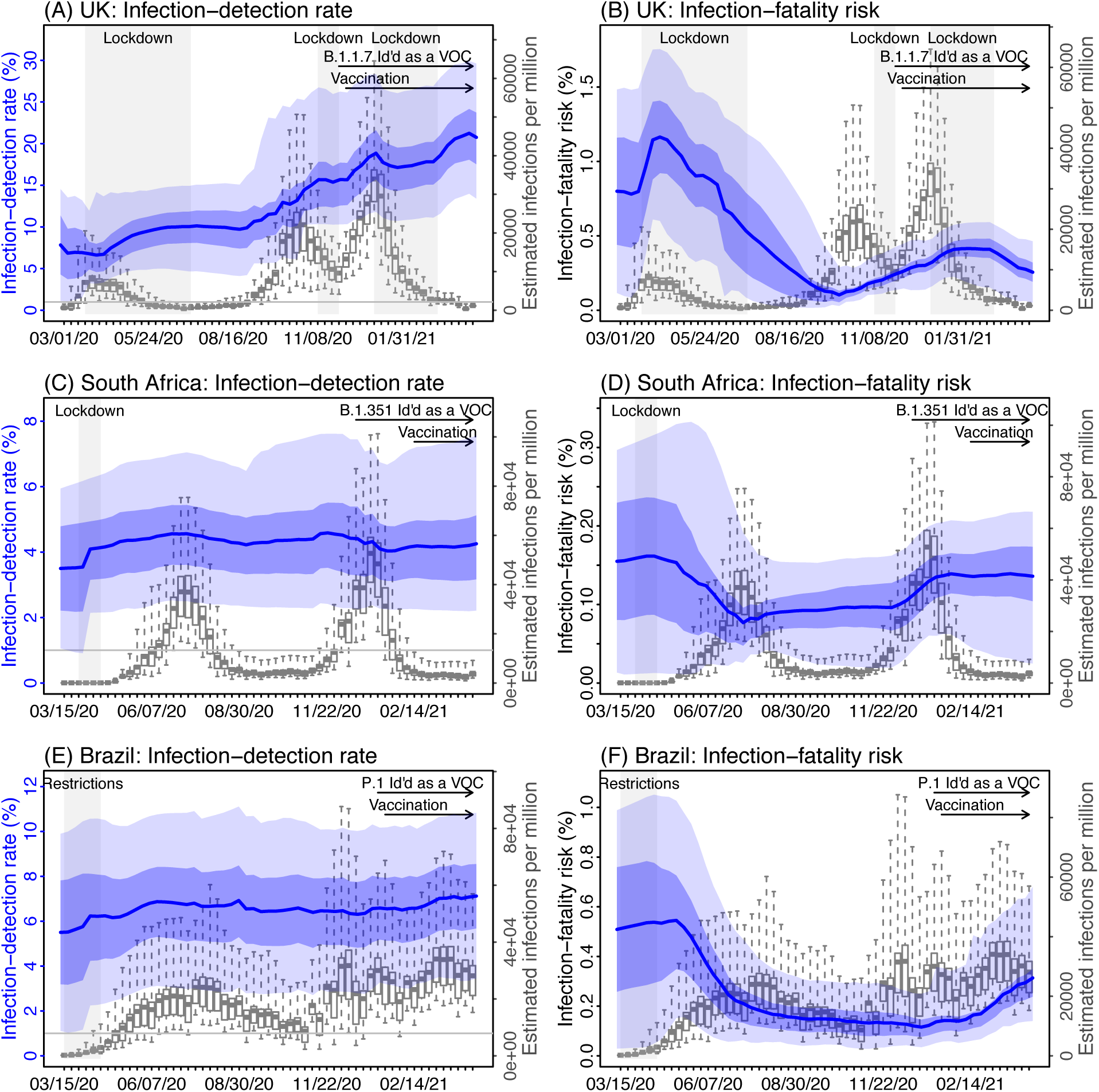
Other model estimates. Left panel shows the estimated infection-detection rate and right panel shows the estimated infection-fatality risk for each week during the study period for the three countries. For comparison, estimated weekly infection rates are superimposed in each plot (right y-axis). Blue lines and surrounding areas show model estimated mean, 50% and 95% CrIs. Boxes and whiskers show model-estimated weekly infection rates (mean, 50% and 95% CrIs). Grey shaded boxes indicate the timing of lockdowns or key period of restrictions; horizontal arrows indicate the timing of variant identification and vaccination rollout.

**Fig S4.**
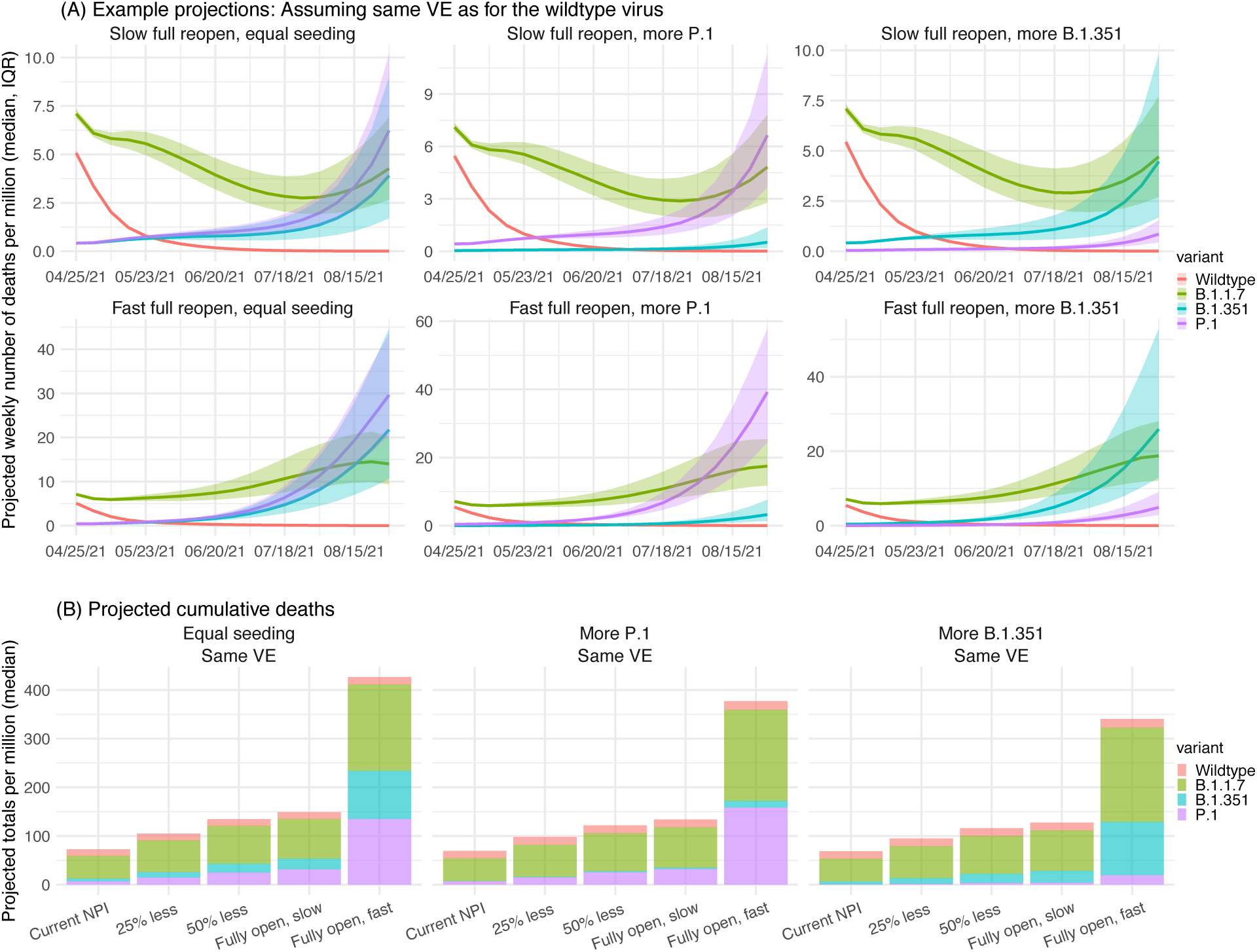
Model projections of *COVID-19 related mortality* under different scenarios of VOC co-circulation and NPI. Due to the uncertainty on the infection fatality risk among breakthrough infections (i.e., those who have been vaccinated), all simulations shown here assume no reduction in vaccine efficacy (VE). All numbers are scaled for 1 million people.

**Fig S5.**
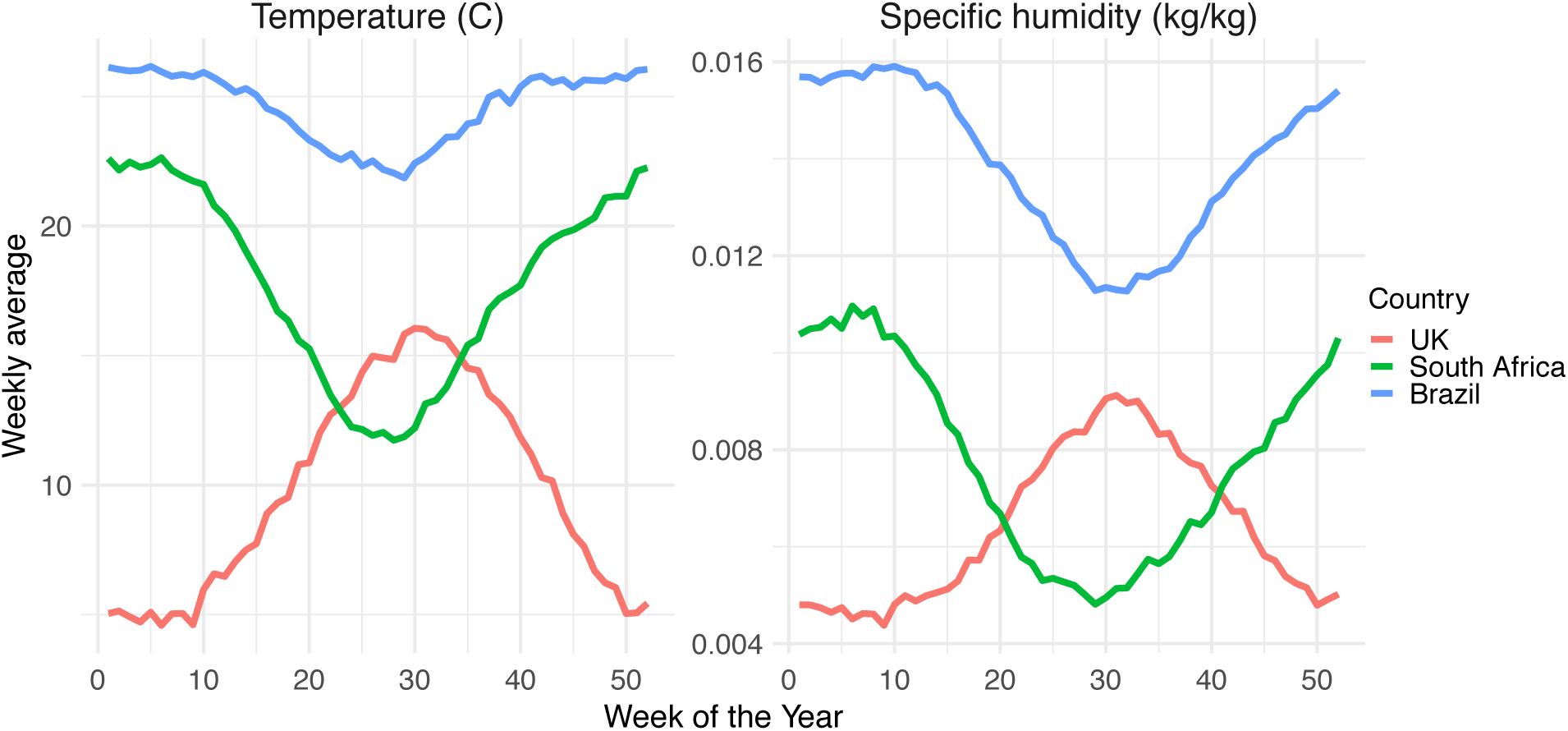
Weekly average temperature and specific humidity for the three countries.

**Table S1.** Model-simulated prevalence of different variants under different scenarios. Numbers show the median (and interquartile range; all in percentage) of tallies over the entire simulation period (i.e. the week of 4/25/2021 to 8/22/2021) for each scenario, as specified in columns 1 (seeding of the B.1.351 and P.1 variant), 2 (NPI), and 3 (reduction on vaccine efficacy).

| Seeding | NPI | VE | Wildtype | B.1.1.7 | B.1.351 | P.1 |
| --- | --- | --- | --- | --- | --- | --- |
| Equal seeding | Current NPI | Same VE | 8.7 (8.1, 9.4) | 65.4 (62.1, 67.9) | 11.9 (9.8, 15) | 13.2 (11.5, 15.2) |
| Equal seeding | Current NPI | High VE | 7.7 (7.1, 8.3) | 63.9 (60.2, 67) | 13.3 (10.5, 17) | 14.3 (12.4, 16.6) |
| Equal seeding | Current NPI | Median VE | 7.6 (6.9, 8.3) | 63.1 (59.5, 66.3) | 13.7 (10.8, 17.6) | 14.6 (12.6, 17) |
| Equal seeding | 25% less | Same VE | 5.7 (4.9, 6.5) | 57.1 (51.4, 62.3) | 15.9 (11.3, 21.8) | 19.5 (15.9, 23.8) |
| Equal seeding | 25% less | High VE | 4.5 (3.8, 5.4) | 52.6 (47, 59) | 19 (12.8, 26.6) | 20.9 (16.9, 26.4) |
| Equal seeding | 25% less | Median VE | 4.3 (3.5, 5.1) | 50.2 (43.1, 56.8) | 20.5 (13.9, 30) | 22.2 (17.6, 27.2) |
| Equal seeding | 50% less | Same VE | 3.8 (3, 4.7) | 48.2 (39.9, 54.8) | 19.5 (12.5, 29.3) | 25.5 (19.7, 32.3) |
| Equal seeding | 50% less | High VE | 2.8 (2.1, 3.5) | 40.7 (32.2, 49.1) | 24.3 (14.7, 37.4) | 27.6 (21.4, 36) |
| Equal seeding | 50% less | Median VE | 2.5 (1.9, 3.3) | 36.6 (28.2, 45) | 27.8 (16.2, 41.2) | 28.7 (21.5, 37.5) |
| Equal seeding | Fully open, slow | Same VE | 2.9 (2.2, 3.7) | 40.5 (32.4, 49.8) | 20.9 (13.1, 35.3) | 28.9 (21.8, 38) |
| Equal seeding | Fully open, slow | High VE | 2 (1.5, 2.7) | 33.9 (25.5, 43.7) | 24.8 (15.1, 41.5) | 32.5 (24, 42.2) |
| Equal seeding | Fully open, slow | Median VE | 1.8 (1.3, 2.4) | 30.6 (22.6, 40) | 30.6 (17, 46.3) | 32.5 (23.1, 42.3) |
| Equal seeding | Fully open, fast | Same VE | 1 (0.9, 1.3) | 26.8 (20.4, 35.4) | 30.1 (17.2, 45.5) | 36.7 (26.3, 48) |
| Equal seeding | Fully open, fast | High VE | 0.8 (0.6, 0.9) | 22.4 (16.7, 30) | 33.7 (20.4, 50.8) | 38.3 (26.7, 50.1) |
| Equal seeding | Fully open, fast | Median VE | 0.7 (0.6, 0.8) | 19.5 (15, 26) | 39.9 (24.1, 56.1) | 36.8 (24.9, 48.9) |
| More P.1 | Current NPI | Same VE | 11.9 (11.1, 12.8) | 71.5 (69.4, 73.5) | 1.6 (1.2, 2.1) | 14.6 (12.9, 16.7) |
| More P.1 | Current NPI | High VE | 10.8 (9.9, 11.6) | 70.9 (68.5, 73.3) | 1.7 (1.3, 2.4) | 16.1 (13.9, 18.6) |
| More P.1 | Current NPI | Median VE | 10.6 (9.8, 11.5) | 70.9 (68.1, 73.1) | 1.9 (1.4, 2.5) | 16.3 (14.1, 18.9) |
| More P.1 | 25% less | Same VE | 8.2 (7.1, 9.4) | 66 (61.6, 69.9) | 2.4 (1.6, 3.5) | 22.4 (18.4, 27.5) |
| More P.1 | 25% less | High VE | 6.9 (5.8, 7.9) | 63.6 (57.7, 68.2) | 2.7 (1.7, 4.4) | 26 (21.2, 31.9) |
| More P.1 | 25% less | Median VE | 6.6 (5.6, 7.5) | 62 (56.4, 66.5) | 3.1 (1.9, 5.1) | 27.3 (22.6, 33) |
| More P.1 | 50% less | Same VE | 5.7 (4.6, 7) | 58.6 (50.8, 64.8) | 3 (1.7, 5) | 31.1 (24.6, 39.6) |
| More P.1 | 50% less | High VE | 4.4 (3.4, 5.4) | 52.5 (44.6, 59.7) | 3.9 (2, 7.5) | 36.5 (29.2, 45.7) |
| More P.1 | 50% less | Median VE | 4.1 (3.2, 5.1) | 49.7 (42.3, 56.9) | 4.5 (2.3, 8.5) | 39.3 (32.1, 47.4) |
| More P.1 | Fully open, slow | Same VE | 4.4 (3.4, 5.8) | 53.3 (44.5, 60.5) | 3.5 (1.8, 6.8) | 36.8 (28.8, 46.1) |
| More P.1 | Fully open, slow | High VE | 3.2 (2.5, 4.2) | 45 (36.8, 53.6) | 4.4 (2.3, 8.8) | 45 (35.7, 53.9) |
| More P.1 | Fully open, slow | Median VE | 3 (2.3, 3.9) | 43 (34.3, 50.8) | 4.9 (2.5, 10.3) | 45.6 (36.1, 55.6) |
| More P.1 | Fully open, fast | Same VE | 1.6 (1.3, 2) | 36.1 (28.6, 45.2) | 5.1 (2.3, 10.8) | 53.2 (43.8, 63.2) |
| More P.1 | Fully open, fast | High VE | 1.2 (1, 1.4) | 30.9 (24.3, 39.2) | 6.9 (3, 14.2) | 56.7 (47.8, 66.2) |
| More P.1 | Fully open, fast | Median VE | 1 (0.9, 1.3) | 27.8 (21.9, 35.4) | 8.4 (3.9, 17.1) | 58.8 (47.7, 67.4) |
| More B.1.351 | Current NPI | Same VE | 12.1 (11.3, 12.9) | 72.5 (69.8, 74.8) | 13.2 (10.9, 16.6) | 1.8 (1.5, 2.2) |
| More B.1.351 | Current NPI | High VE | 10.9 (10, 11.8) | 72.2 (68.7, 74.9) | 14.6 (11.5, 18.6) | 1.9 (1.5, 2.4) |
| More B.1.351 | Current NPI | Median VE | 10.6 (9.8, 11.5) | 71.5 (67.6, 74.6) | 15.5 (12.3, 19.8) | 2 (1.6, 2.5) |
| More B.1.351 | 25% less | Same VE | 8.4 (7.2, 9.6) | 68.9 (62, 73.6) | 19.3 (13.7, 27) | 2.8 (2, 3.8) |
| More B.1.351 | 25% less | High VE | 7 (5.9, 8.1) | 66.9 (58.1, 72.6) | 22.3 (15.6, 32.3) | 3.2 (2.3, 4.5) |
| More B.1.351 | 25% less | Median VE | 6.5 (5.4, 7.8) | 62.9 (53.8, 70.5) | 26.3 (17.5, 36.5) | 3.3 (2.4, 4.7) |
| More B.1.351 | 50% less | Same VE | 5.9 (4.7, 7.3) | 62.7 (51.1, 70.5) | 26.4 (17.1, 39.2) | 4.1 (2.7, 6) |
| More B.1.351 | 50% less | High VE | 4.5 (3.5, 5.8) | 56.6 (43.7, 67.4) | 32.4 (20.1, 46.9) | 4.9 (3.3, 7.3) |
| More B.1.351 | 50% less | Median VE | 4.1 (3, 5.3) | 53 (39.6, 63.4) | 36.8 (24.3, 52.5) | 5 (3.2, 7.6) |
| More B.1.351 | Fully open, slow | Same VE | 4.7 (3.5, 6.2) | 58.2 (43.8, 68.5) | 30.7 (18, 47) | 4.9 (3.2, 7.6) |
| More B.1.351 | Fully open, slow | High VE | 3.4 (2.4, 4.6) | 50 (35.9, 62.4) | 38.5 (23.5, 56.1) | 5.7 (3.5, 8.9) |
| More B.1.351 | Fully open, slow | Median VE | 3.1 (2.2, 4.2) | 44.7 (30.9, 57.4) | 43.6 (28.2, 61) | 5.9 (3.3, 9.5) |
| More B.1.351 | Fully open, fast | Same VE | 1.7 (1.3, 2.3) | 42.2 (28.6, 55.5) | 44.4 (28.7, 63.1) | 7.5 (4.4, 13.1) |
| More B.1.351 | Fully open, fast | High VE | 1.2 (1, 1.6) | 34.4 (23, 48.2) | 53.4 (35.1, 69.1) | 8.2 (4.7, 13.7) |
| More B.1.351 | Fully open, fast | Median VE | 1.1 (0.8, 1.4) | 29.2 (19.9, 41.9) | 59.7 (42.1, 72.6) | 7.7 (4.3, 13.2) |

**Table S2.**
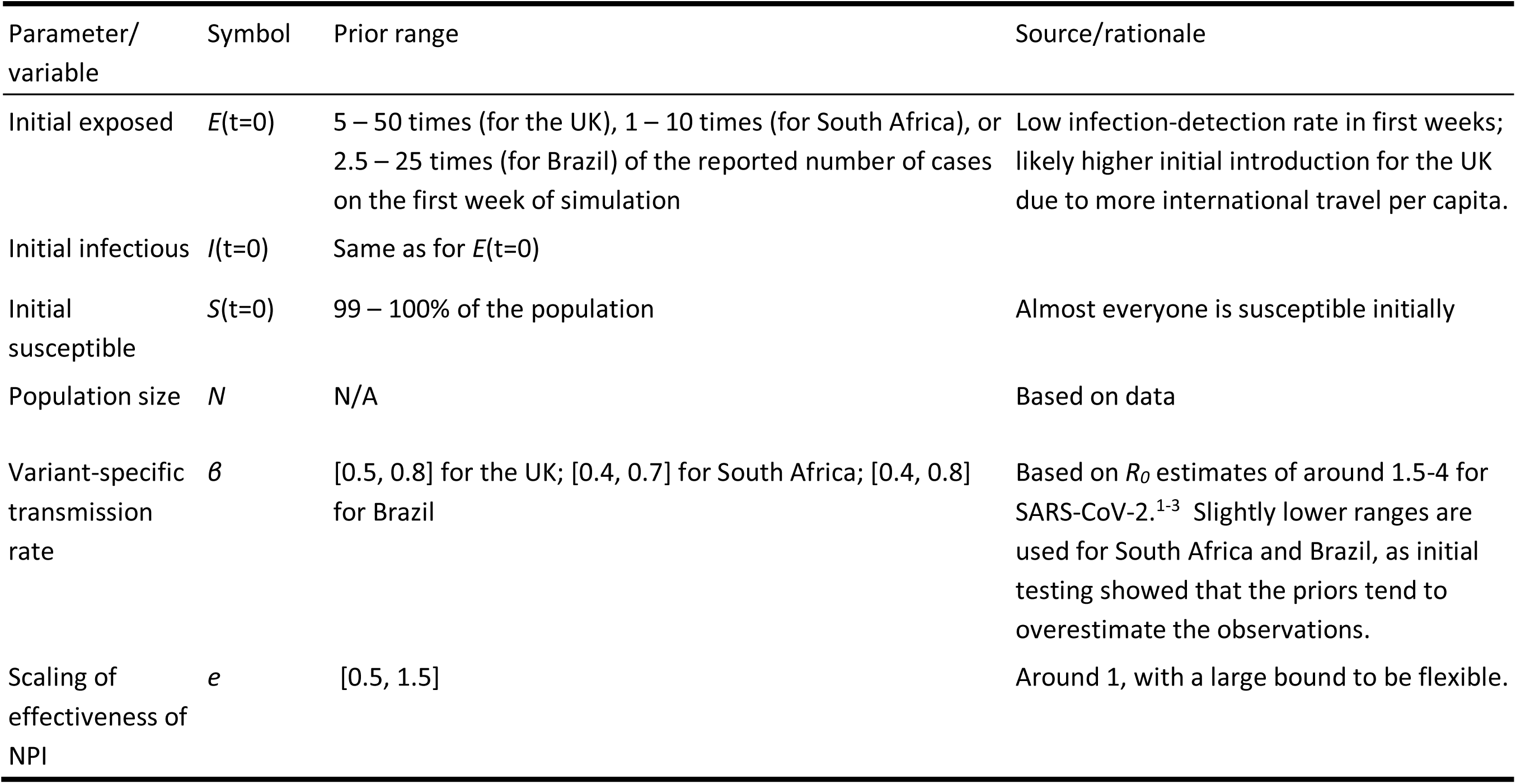
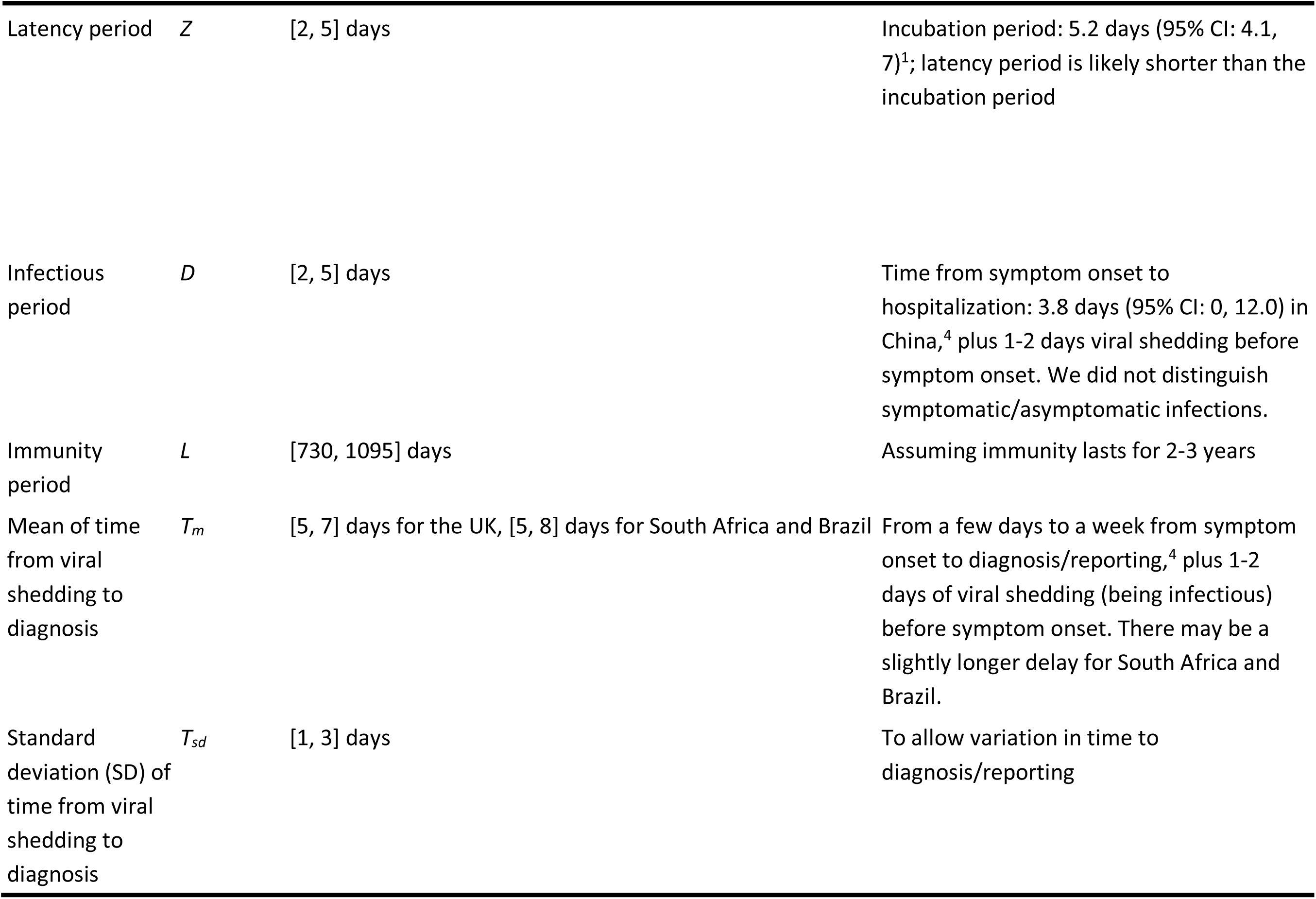
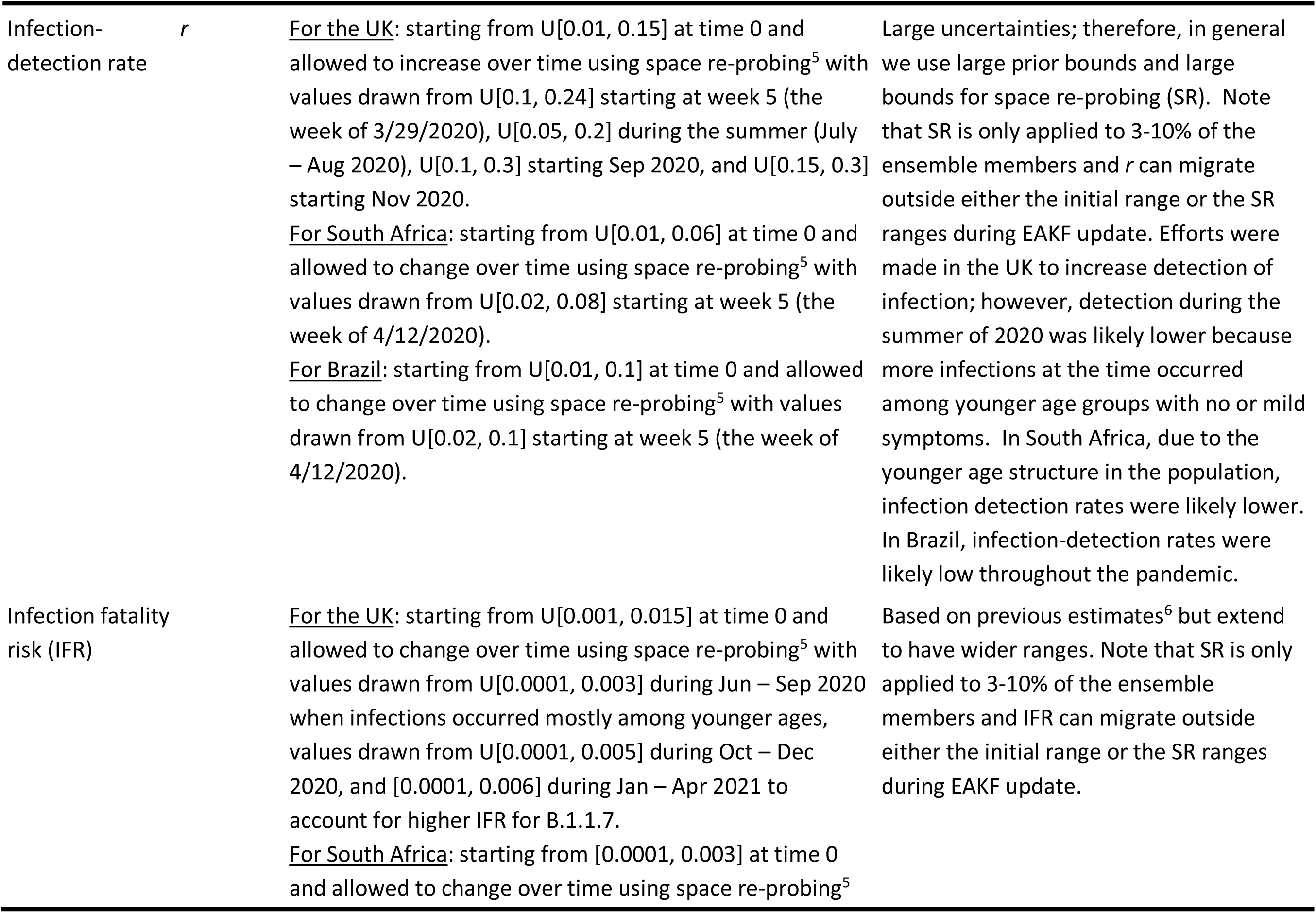
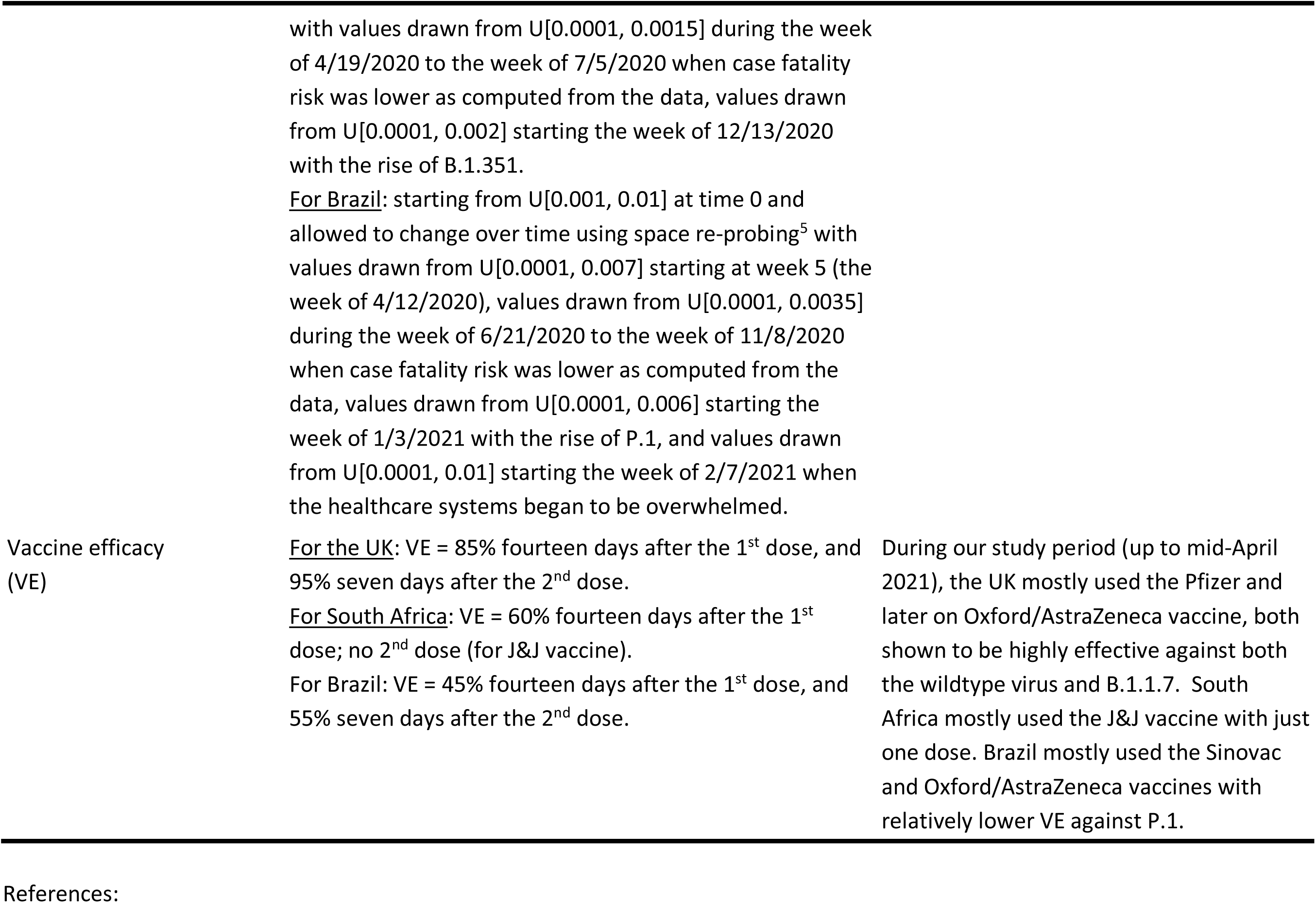
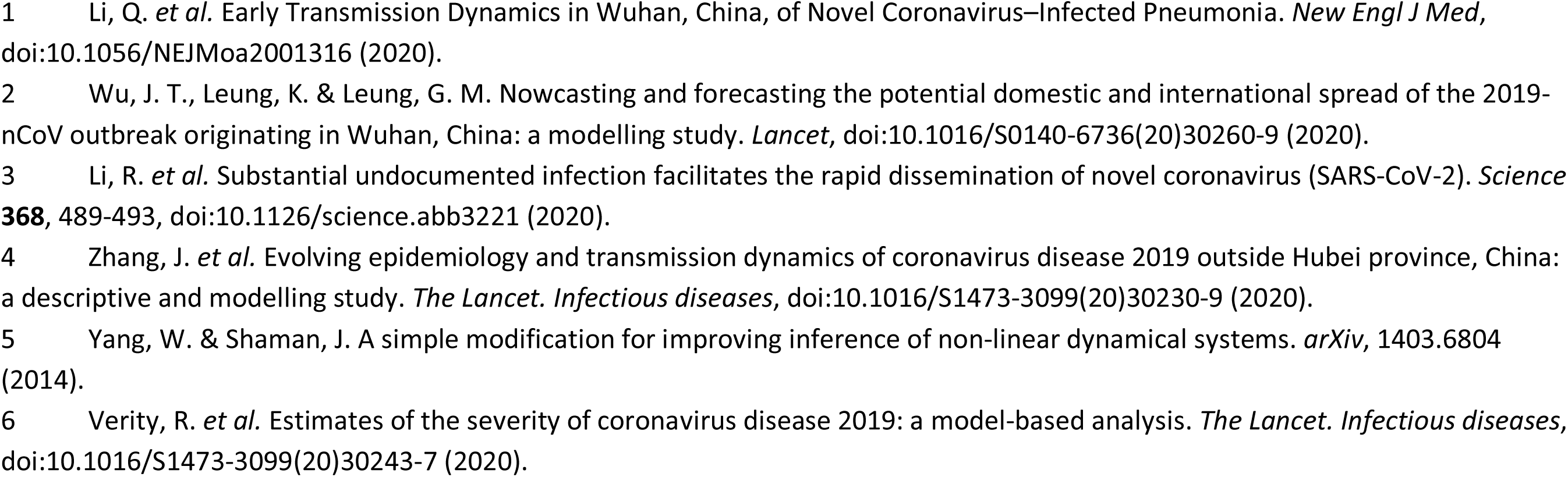
Prior ranges for the parameters used in the model-inference system for the three countries. Parameter/ Symbol Prior range Source/rationale

**Table S3.** Parameters used to generate the synthetic data for model validation Truth Settings

| Truth | Settings |
| --- | --- |
| Wildtype-virus,<br>same for<br>all truths | $\beta = 0.65$ per day; $Z = 3.5$ days; $D = 3.5$ days; $L = 2.5$ years; $T_m = 6$ days; $T_{sd} = 2$ days; IFR = 0.7% |
| Variant in<br>Truth 1 | Large outbreak during the first wave (See Figs 1 and S1). No increase in transmissibility (i.e. same $\beta$ and $D$ as for the wildtype virus); 80% increase in immune escape. Other parameters same as the wildtype virus. |
| Variant in<br>Truth 2 | Large outbreak during the first wave (See Figs 1 and S1). 50% increase in transmissibility (i.e. $\beta = 0.65 \times 1.5 = 0.975$ per day); no immune escape. Other parameters same as the wildtype virus. |
| Variant in<br>Truth 3 | Large outbreak during the first wave (See Figs 1 and S1). 50% increase in transmissibility (i.e. $\beta = 0.65 \times 1.5 = 0.975$ per day); 80% increase in immune escape. Other parameters same as the wildtype virus. |
| Variant in<br>Truth 4 | Large outbreak during the first wave (See Figs 1 and S1). 25% increase in transmissibility (i.e. $\beta = 0.65 \times 1.25 = 0.8125$ per day); 40% increase in immune escape. Other parameters same as the wildtype virus. |
| Variant in<br>Truth 5 | Small outbreak during the first wave (See Figs 1 and S1). 50% increase in transmissibility (i.e. $\beta = 0.65 \times 1.5 = 0.975$ per day); no immune escape. Other parameters same as the wildtype virus. |
| Infection-detection<br>rate | 20% for results shown in Fig 1 and 10% for results shown in Fig S1. |

**Table S4.** Cumulative vaccination coverage used in the multi-variant, age-structured model simulations. Baseline vaccination coverage (as of 4/24/2021) is based on data for NYC. Projected vaccination rates for the simulation period (from the week starting 4/25/2021 to the week ending 8/29/2021) are based on data 10 days preceding the simulation and assuming a cumulative vaccination uptake of 80%. Note that ages under 15 (i.e., <1, 1-4, 5-14 years) are combined in the same column as they were not eligible to receive the vaccines at the time of this study. In addition, the numbers are aggregated from neighborhood-level estimates and thus could slightly exceed the 80% total for some age groups when some neighborhoods had actual vaccination coverage above 80% at baseline.

| Time point | Vaccine dose | Vaccination coverage (%) by age group (in year) |  |  |  |  |  |
| --- | --- | --- | --- | --- | --- | --- | --- |
|  |  | 0-14 | 15-24 | 25-44 | 45-64 | 65-74 | 75+ |
| Start of simulation: | 1st dose | 0 | 25.4 | 41.7 | 52.9 | 69 | 58.2 |
| 4/24/2021 | 2nd dose | 0 | 10.5 | 26.7 | 40.7 | 62.6 | 53.6 |
| End of simulation: | 1st dose | 0 | 70 | 76 | 79.1 | 80.6 | 76.8 |
| 8/29/2021 | 2nd dose | 0 | 65.4 | 73.4 | 78.7 | 80.3 | 74.5 |

**Table S5.**
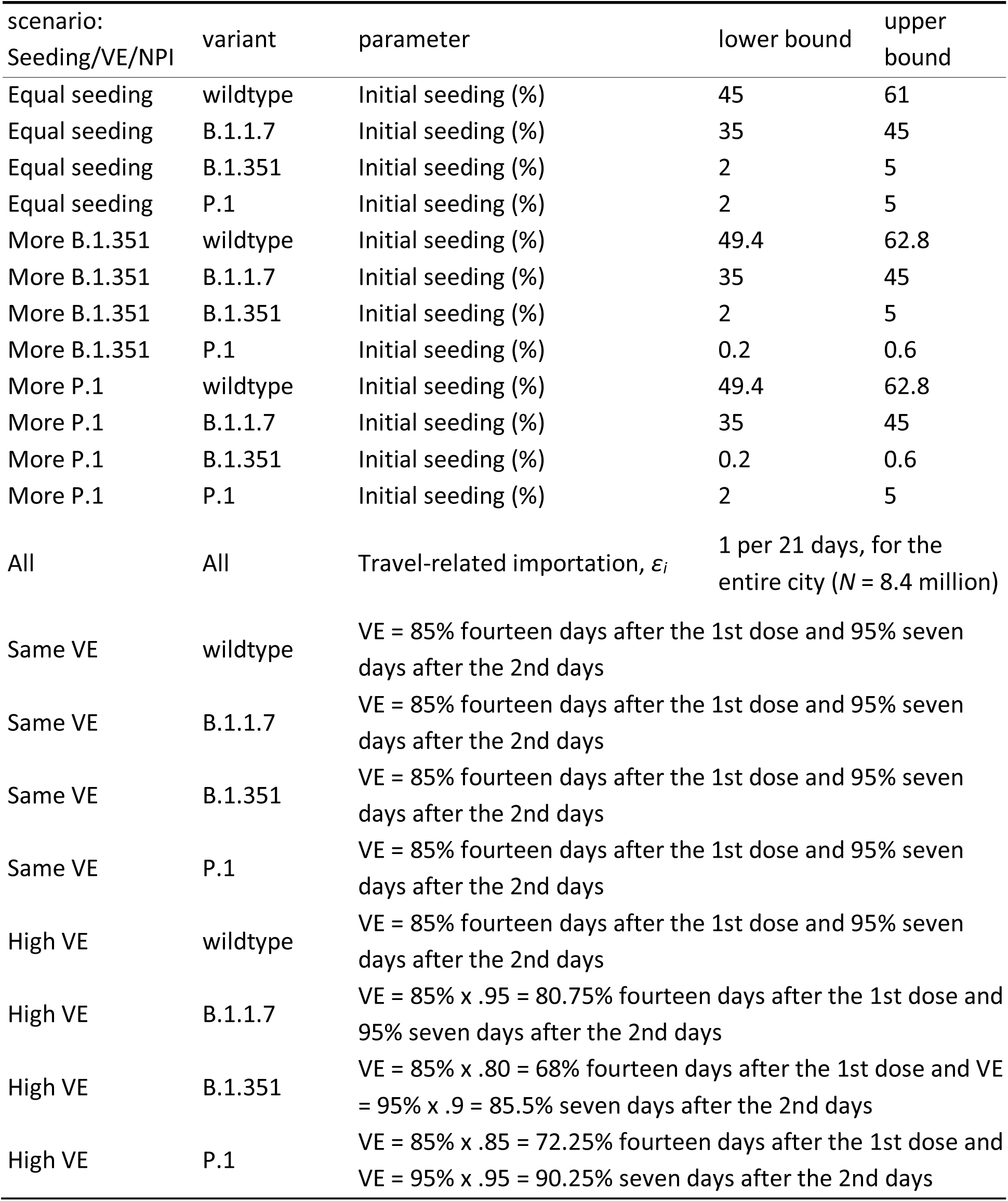
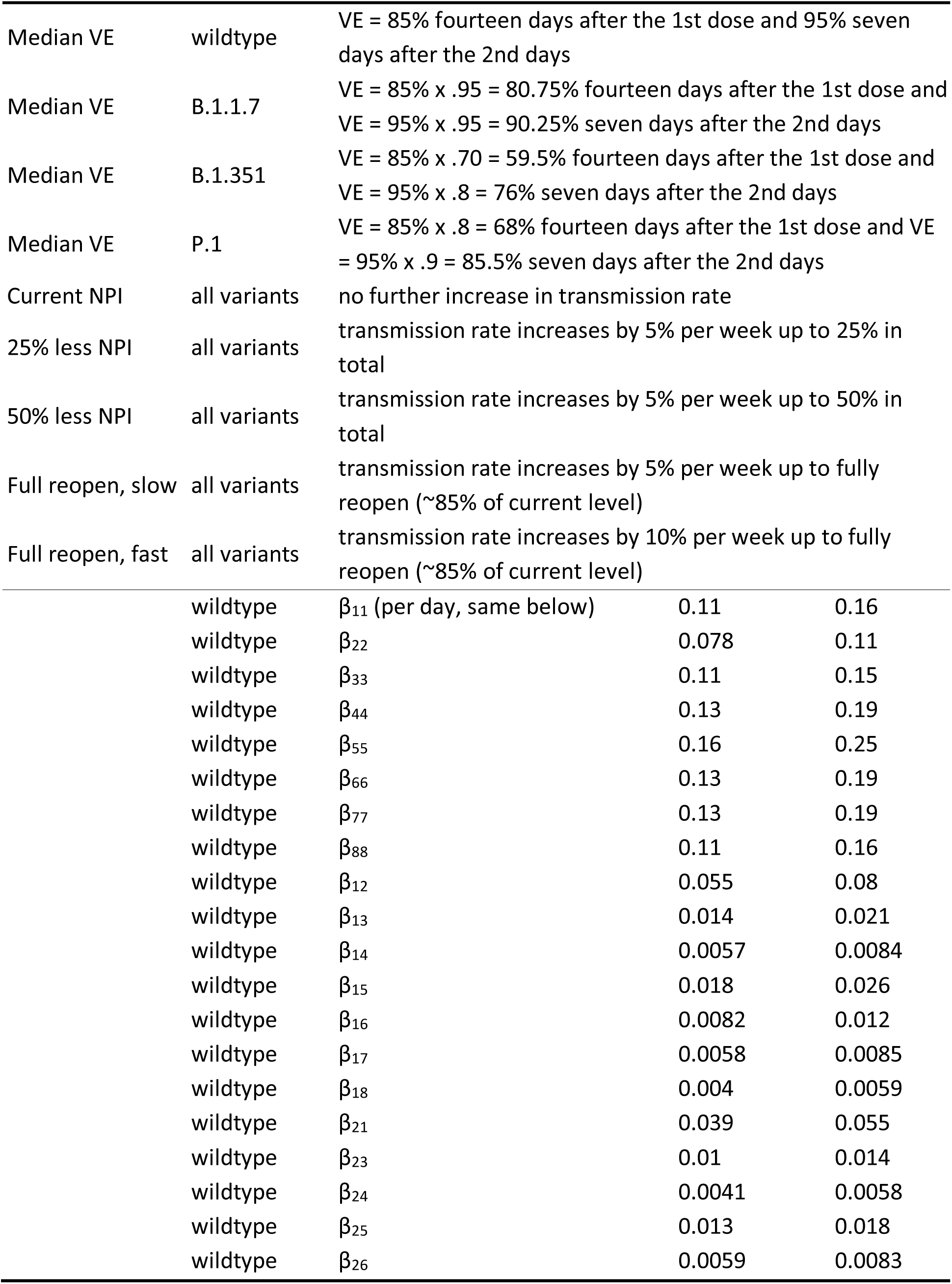
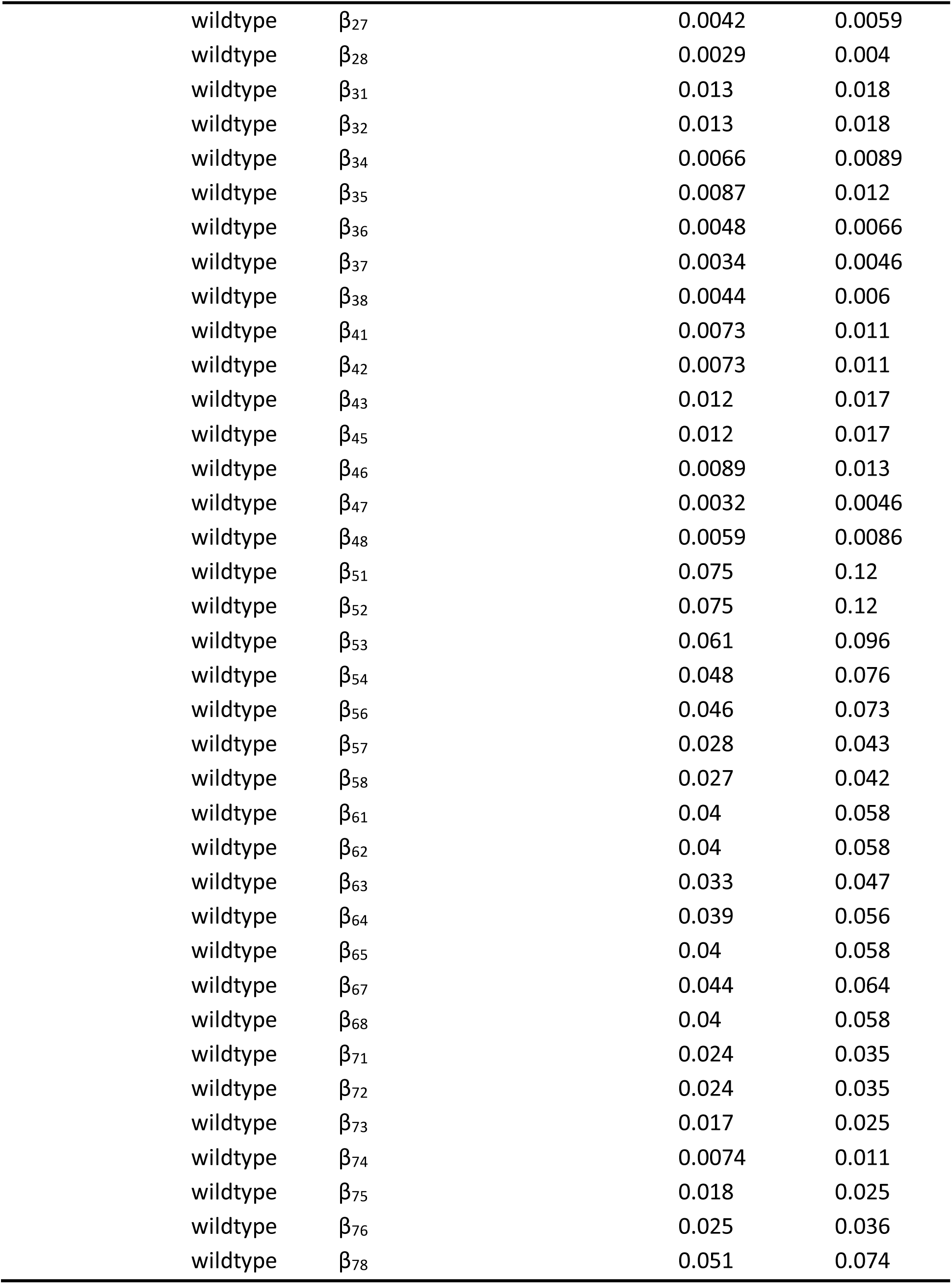
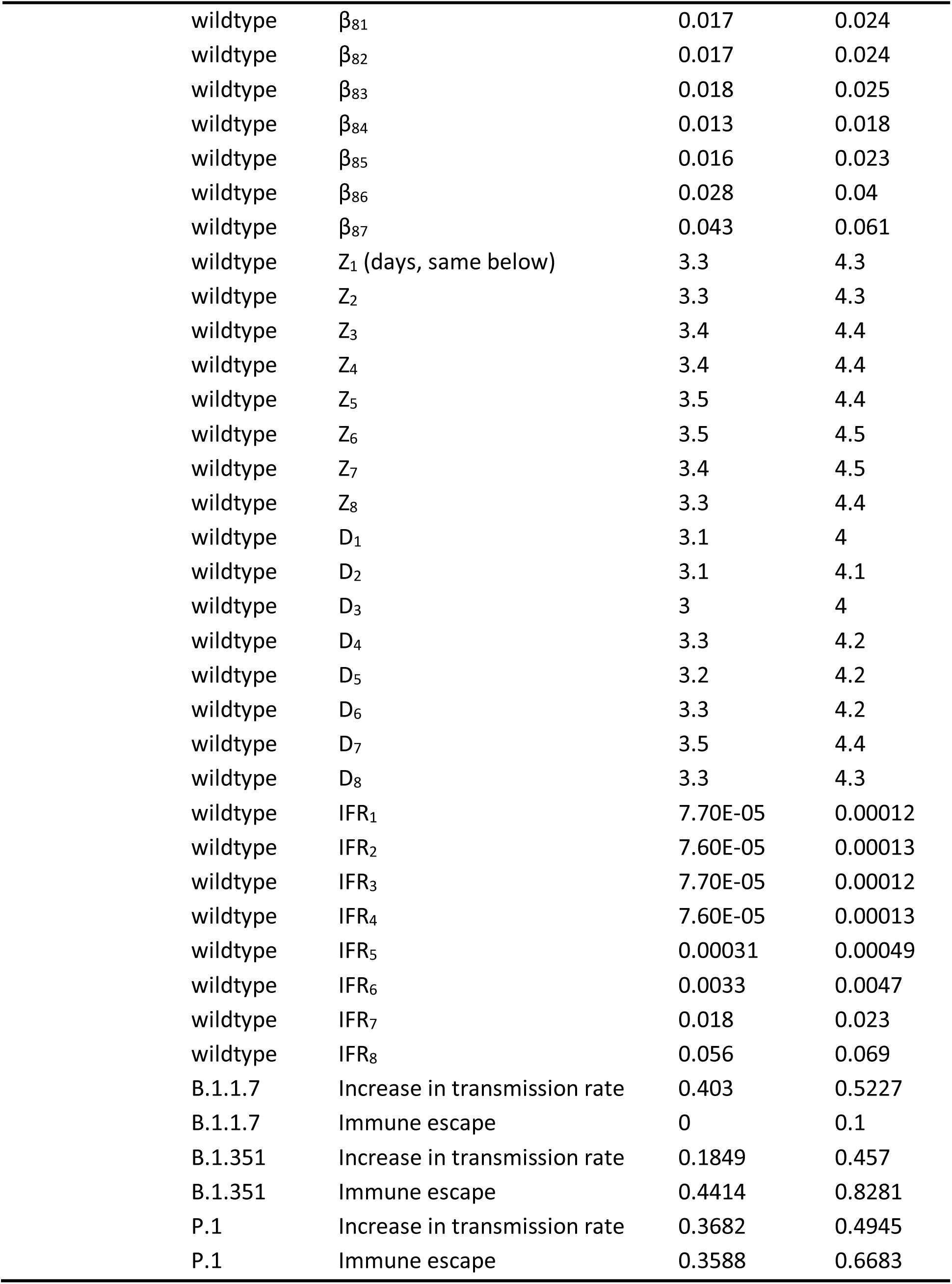
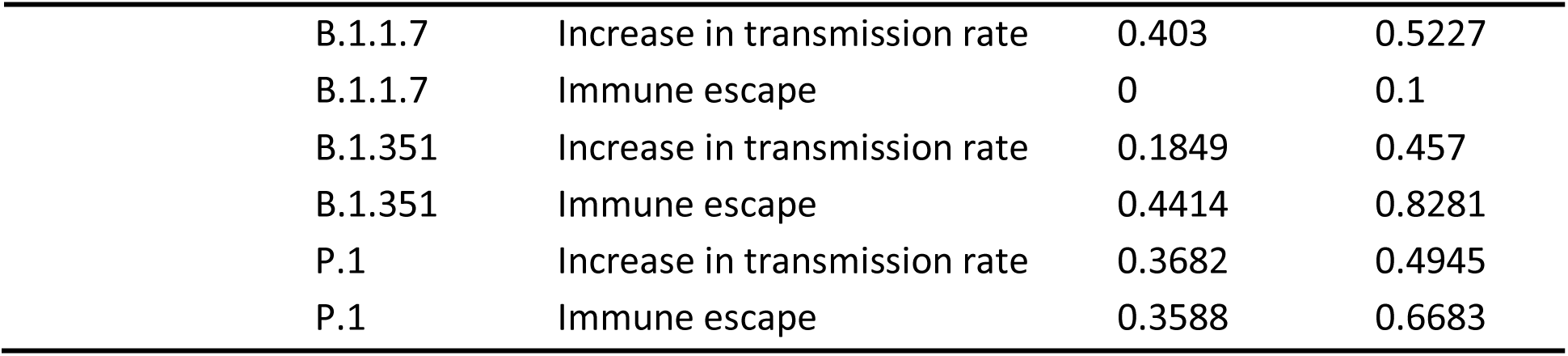
Parameter settings for different scenarios to simulate and project the impact of different variants of concern. Initial conditions and parameters are randomly drawn from uniform distributions with lower and upper bounds as specified below, based on estimates for NYC made for the week of 4/18/2021 using data during 3/1/2020 – 4/24/2021. Numbers associated with the parameter names denote the corresponding age groups.

| scenario:<br>Seeding/VE/NPI | variant | parameter | lower bound | upper bound |
| --- | --- | --- | --- | --- |
| Equal seeding | wildtype | Initial seeding (%) | 45 | 61 |
| Equal seeding | B.1.1.7 | Initial seeding (%) | 35 | 45 |
| Equal seeding | B.1.351 | Initial seeding (%) | 2 | 5 |
| Equal seeding | P.1 | Initial seeding (%) | 2 | 5 |
| More B.1.351 | wildtype | Initial seeding (%) | 49.4 | 62.8 |
| More B.1.351 | B.1.1.7 | Initial seeding (%) | 35 | 45 |
| More B.1.351 | B.1.351 | Initial seeding (%) | 2 | 5 |
| More B.1.351 | P.1 | Initial seeding (%) | 0.2 | 0.6 |
| More P.1 | wildtype | Initial seeding (%) | 49.4 | 62.8 |
| More P.1 | B.1.1.7 | Initial seeding (%) | 35 | 45 |
| More P.1 | B.1.351 | Initial seeding (%) | 0.2 | 0.6 |
| More P.1 | P.1 | Initial seeding (%) | 2 | 5 |
| All | All | Travel-related importation, $\varepsilon_i$ | 1 per 21 days, for the entire city ($N = 8.4$ million) | |
| Same VE | wildtype | VE = 85% fourteen days after the 1st dose and 95% seven days after the 2nd days |  |  |
| Same VE | B.1.1.7 | VE = 85% fourteen days after the 1st dose and 95% seven days after the 2nd days |  |  |
| Same VE | B.1.351 | VE = 85% fourteen days after the 1st dose and 95% seven days after the 2nd days |  |  |
| Same VE | P.1 | VE = 85% fourteen days after the 1st dose and 95% seven days after the 2nd days |  |  |
| High VE | wildtype | VE = 85% fourteen days after the 1st dose and 95% seven days after the 2nd days |  |  |
| High VE | B.1.1.7 | VE = 85% x .95 = 80.75% fourteen days after the 1st dose and 95% seven days after the 2nd days |  |  |
| High VE | B.1.351 | VE = 85% x .80 = 68% fourteen days after the 1st dose and VE = 95% x .9 = 85.5% seven days after the 2nd days |  |  |
| High VE | P.1 | VE = 85% x .85 = 72.25% fourteen days after the 1st dose and VE = 95% x .95 = 90.25% seven days after the 2nd days |  |  |
| Median VE | wildtype | VE = 85% fourteen days after the 1st dose and 95% seven days after the 2nd days |  |  |
| Median VE | B.1.1.7 | VE = 85% x .95 = 80.75% fourteen days after the 1st dose and<br>VE = 95% x .95 = 90.25% seven days after the 2nd days |  |  |
| Median VE | B.1.351 | VE = 85% x .70 = 59.5% fourteen days after the 1st dose and<br>VE = 95% x .8 = 76% seven days after the 2nd days |  |  |
| Median VE | P.1 | VE = 85% x .8 = 68% fourteen days after the 1st dose and VE<br>= 95% x .9 = 85.5% seven days after the 2nd days |  |  |
| Current NPI | all variants | no further increase in transmission rate |  |  |
| 25% less NPI | all variants | transmission rate increases by 5% per week up to 25% in total |  |  |
| 50% less NPI | all variants | transmission rate increases by 5% per week up to 50% in total |  |  |
| Full reopen, slow | all variants | transmission rate increases by 5% per week up to fully reopen (~85% of current level) |  |  |
| Full reopen, fast | all variants | transmission rate increases by 10% per week up to fully reopen (~85% of current level) |  |  |
| | wildtype | $\beta_{11}$ (per day, same below) | 0.11 | 0.16 |
| | wildtype | $\beta_{22}$ | 0.078 | 0.11 |
| | wildtype | $\beta_{33}$ | 0.11 | 0.15 |
| | wildtype | $\beta_{44}$ | 0.13 | 0.19 |
| | wildtype | $\beta_{55}$ | 0.16 | 0.25 |
| | wildtype | $\beta_{66}$ | 0.13 | 0.19 |
| | wildtype | $\beta_{77}$ | 0.13 | 0.19 |
| | wildtype | $\beta_{88}$ | 0.11 | 0.16 |
| | wildtype | $\beta_{12}$ | 0.055 | 0.08 |
| | wildtype | $\beta_{13}$ | 0.014 | 0.021 |
| | wildtype | $\beta_{14}$ | 0.0057 | 0.0084 |
| | wildtype | $\beta_{15}$ | 0.018 | 0.026 |
| | wildtype | $\beta_{16}$ | 0.0082 | 0.012 |
| | wildtype | $\beta_{17}$ | 0.0058 | 0.0085 |
| | wildtype | $\beta_{18}$ | 0.004 | 0.0059 |
| | wildtype | $\beta_{21}$ | 0.039 | 0.055 |
| | wildtype | $\beta_{23}$ | 0.01 | 0.014 |
| | wildtype | $\beta_{24}$ | 0.0041 | 0.0058 |
| | wildtype | $\beta_{25}$ | 0.013 | 0.018 |
| | wildtype | $\beta_{26}$ | 0.0059 | 0.0083 |

|  |  |  |  |
| --- | --- | --- | --- |
| wildtype | $\beta_{27}$ | 0.0042 | 0.0059 |
| wildtype | $\beta_{28}$ | 0.0029 | 0.004 |
| wildtype | $\beta_{31}$ | 0.013 | 0.018 |
| wildtype | $\beta_{32}$ | 0.013 | 0.018 |
| wildtype | $\beta_{34}$ | 0.0066 | 0.0089 |
| wildtype | $\beta_{35}$ | 0.0087 | 0.012 |
| wildtype | $\beta_{36}$ | 0.0048 | 0.0066 |
| wildtype | $\beta_{37}$ | 0.0034 | 0.0046 |
| wildtype | $\beta_{38}$ | 0.0044 | 0.006 |
| wildtype | $\beta_{41}$ | 0.0073 | 0.011 |
| wildtype | $\beta_{42}$ | 0.0073 | 0.011 |
| wildtype | $\beta_{43}$ | 0.012 | 0.017 |
| wildtype | $\beta_{45}$ | 0.012 | 0.017 |
| wildtype | $\beta_{46}$ | 0.0089 | 0.013 |
| wildtype | $\beta_{47}$ | 0.0032 | 0.0046 |
| wildtype | $\beta_{48}$ | 0.0059 | 0.0086 |
| wildtype | $\beta_{51}$ | 0.075 | 0.12 |
| wildtype | $\beta_{52}$ | 0.075 | 0.12 |
| wildtype | $\beta_{53}$ | 0.061 | 0.096 |
| wildtype | $\beta_{54}$ | 0.048 | 0.076 |
| wildtype | $\beta_{56}$ | 0.046 | 0.073 |
| wildtype | $\beta_{57}$ | 0.028 | 0.043 |
| wildtype | $\beta_{58}$ | 0.027 | 0.042 |
| wildtype | $\beta_{61}$ | 0.04 | 0.058 |
| wildtype | $\beta_{62}$ | 0.04 | 0.058 |
| wildtype | $\beta_{63}$ | 0.033 | 0.047 |
| wildtype | $\beta_{64}$ | 0.039 | 0.056 |
| wildtype | $\beta_{65}$ | 0.04 | 0.058 |
| wildtype | $\beta_{67}$ | 0.044 | 0.064 |
| wildtype | $\beta_{68}$ | 0.04 | 0.058 |
| wildtype | $\beta_{71}$ | 0.024 | 0.035 |
| wildtype | $\beta_{72}$ | 0.024 | 0.035 |
| wildtype | $\beta_{73}$ | 0.017 | 0.025 |
| wildtype | $\beta_{74}$ | 0.0074 | 0.011 |
| wildtype | $\beta_{75}$ | 0.018 | 0.025 |
| wildtype | $\beta_{76}$ | 0.025 | 0.036 |
| wildtype | $\beta_{78}$ | 0.051 | 0.074 |

|  |  |  |  |
| --- | --- | --- | --- |
| wildtype | $\beta_{81}$ | 0.017 | 0.024 |
| wildtype | $\beta_{82}$ | 0.017 | 0.024 |
| wildtype | $\beta_{83}$ | 0.018 | 0.025 |
| wildtype | $\beta_{84}$ | 0.013 | 0.018 |
| wildtype | $\beta_{85}$ | 0.016 | 0.023 |
| wildtype | $\beta_{86}$ | 0.028 | 0.04 |
| wildtype | $\beta_{87}$ | 0.043 | 0.061 |
| wildtype | $Z_1$ (days, same below) | 3.3 | 4.3 |
| wildtype | $Z_2$ | 3.3 | 4.3 |
| wildtype | $Z_3$ | 3.4 | 4.4 |
| wildtype | $Z_4$ | 3.4 | 4.4 |
| wildtype | $Z_5$ | 3.5 | 4.4 |
| wildtype | $Z_6$ | 3.5 | 4.5 |
| wildtype | $Z_7$ | 3.4 | 4.5 |
| wildtype | $Z_8$ | 3.3 | 4.4 |
| wildtype | $D_1$ | 3.1 | 4 |
| wildtype | $D_2$ | 3.1 | 4.1 |
| wildtype | $D_3$ | 3 | 4 |
| wildtype | $D_4$ | 3.3 | 4.2 |
| wildtype | $D_5$ | 3.2 | 4.2 |
| wildtype | $D_6$ | 3.3 | 4.2 |
| wildtype | $D_7$ | 3.5 | 4.4 |
| wildtype | $D_8$ | 3.3 | 4.3 |
| wildtype | $IFR_1$ | 7.70E-05 | 0.00012 |
| wildtype | $IFR_2$ | 7.60E-05 | 0.00013 |
| wildtype | $IFR_3$ | 7.70E-05 | 0.00012 |
| wildtype | $IFR_4$ | 7.60E-05 | 0.00013 |
| wildtype | $IFR_5$ | 0.00031 | 0.00049 |
| wildtype | $IFR_6$ | 0.0033 | 0.0047 |
| wildtype | $IFR_7$ | 0.018 | 0.023 |
| wildtype | $IFR_8$ | 0.056 | 0.069 |
| B.1.1.7 | Increase in transmission rate | 0.403 | 0.5227 |
| B.1.1.7 | Immune escape | 0 | 0.1 |
| B.1.351 | Increase in transmission rate | 0.1849 | 0.457 |
| B.1.351 | Immune escape | 0.4414 | 0.8281 |
| P.1 | Increase in transmission rate | 0.3682 | 0.4945 |
| P.1 | Immune escape | 0.3588 | 0.6683 |
| B.1.1.7 | Increase in transmission rate | 0.403 | 0.5227 |
| B.1.1.7 | Immune escape | 0 | 0.1 |
| B.1.351 | Increase in transmission rate | 0.1849 | 0.457 |
| B.1.351 | Immune escape | 0.4414 | 0.8281 |
| P.1 | Increase in transmission rate | 0.3682 | 0.4945 |
| P.1 | Immune escape | 0.3588 | 0.6683 |

